# Leveraging Hand-Crafted Radiomics on Multicenter FLAIR MRI for Predicting Disability Progression in People with Multiple Sclerosis

**DOI:** 10.1101/2025.01.23.25320971

**Authors:** Hamza Khan, Henry C Woodruff, Diana L. Giraldo, Lorin Werthen-Brabants, Shruti Atul Mali, Sina Amirrajab, Edward De Brouwer, Veronica Popescu, Bart Van Wijmeersch, Oliver Gerlach, Jan Sijbers, Liesbet M. Peeters, Philippe Lambin

**Author notes:** Corresponding author Philippe Lambin. These authors have contributed equally to this work (Henry C Woodruff, Diana L. Giraldo, Liesbet M. Peeters, Philippe Lambin).

## Abstract

Multiple sclerosis (MS) is a chronic autoimmune disease of the central nervous system that results in varying degrees of functional impairment. Conventional tools, such as the Expanded Disability Status Scale (EDSS), lack sensitivity to subtle changes in disease progression. Radiomics offers a quantitative imaging approach to address this limitation. This study used machine learning (ML) and radiomics features derived from T2-weighted Fluid-Attenuated Inversion Recovery (FLAIR) magnetic resonance images (MRI) to predict disability progression in people with MS (PwMS).

A retrospective analysis was performed on real-world data from 247 PwMS across two centers. Disability progression was defined using EDSS changes over two years. FLAIR MRIs were preprocessed using bias-field correction, intensity normalisation, and super-resolution reconstruction for low-resolution images. White matter lesions (WML) were segmented using the Lesion Segmentation Toolbox (LST), and MRI tissue segmentation was performed using sequence Adaptive Multimodal SEGmentation. Radiomics features from WML and normal-appearing white matter (NAWM) were extracted using PyRadiomics, harmonised with Longitudinal ComBat, and reduced via Spearman correlation and recursive feature elimination. Elastic Net, Balanced Random Forest (BRFC), and Light Gradient-Boosting Machine (LGBM) models were evaluated on validation data and subsequently tested on unseen data.

The LGBM model with harmonised radiomics and clinical features outperformed the clinical only model by achieving a test performance of PR AUC of 0.20 and a ROC AUC of 0.64. Key predictive features, among others, included GLCM maximum probability (WML), GLDM dependence non uniformity (NAWM). Short-term changes (longitudinal imaging approach) showed limited predictive power by achieving a PR AUC of 0.11 and a ROC AUC of 0.69.

These findings support the use of ML models trained on radiomics features integrated with clinical data for predicting disability progression in PwMS. Future studies should validate these findings in larger, balanced datasets and explore advanced approaches, such as deep learning and foundation models, to enhance predictive performance.

## Introduction

Multiple Sclerosis (MS) is a chronic neuroinflammatory autoimmune disease of the central nervous system (CNS) characterised by demyelination and axonal damage, resulting in varying degrees of loss of functional capabilities (Calabresi, 2004). As of 2024, around 2.9 million people with MS (PwMS) exist worldwide (*Number of People with MS | Atlas of MS*, n.d.). Common symptoms include loss of sensation and motor, autonomic, and neurocognitive dysfunction, although the nature of the symptoms depends on the site of the lesions in the CNS as well as other underlying factors such as age, gender, comorbidities, and different biological mechanisms responsible for inflammation and neurodegeneration (Sospedra & Martin, 2005).

Most PwMS experience disease onset at a relatively young age (Walton et al., 2020). Since no cure for MS exists (Ferrè et al., 2023), early diagnosis coupled with the initiation of disease-modifying therapy in a timely fashion, can alter the clinical course of MS (Edinger & Habibi, 2024) and thus impede neurological damage and improve the quality of life of PwMS (Noyes & Weinstock-Guttman, 2013). However, due to the heterogeneity in MS progression and the incomplete understanding of its pathophysiology, predicting long-term disability remains challenging and can make it difficult for healthcare professionals to diagnose MS earlier and predict prognosis adequately (Thompson et al., 2018; Tilling et al., 2016). The ability to predict long-term disability progression in MS has critical clinical implications. Reliable predictions can help healthcare professionals adapt the frequency and intensity of follow-ups, guide the course of disease-modifying therapies, and enable better stratification of patients for clinical trials. By identifying patients at higher risk of progression, clinicians could focus resources more effectively, improve patient outcomes, and reduce the burden on healthcare systems. Furthermore, accurate prognostic tools would help researchers design clinical trials with more homogenous cohorts, improving the statistical power to detect treatment effects and accelerating the development of new therapies (Dennison et al., 2018; Inojosa et al., 2021). Figure 1 illustrates magnetic resonance imaging (MRI) scans of clinically progressive and non progressive PwMS.

**Figure 1:**
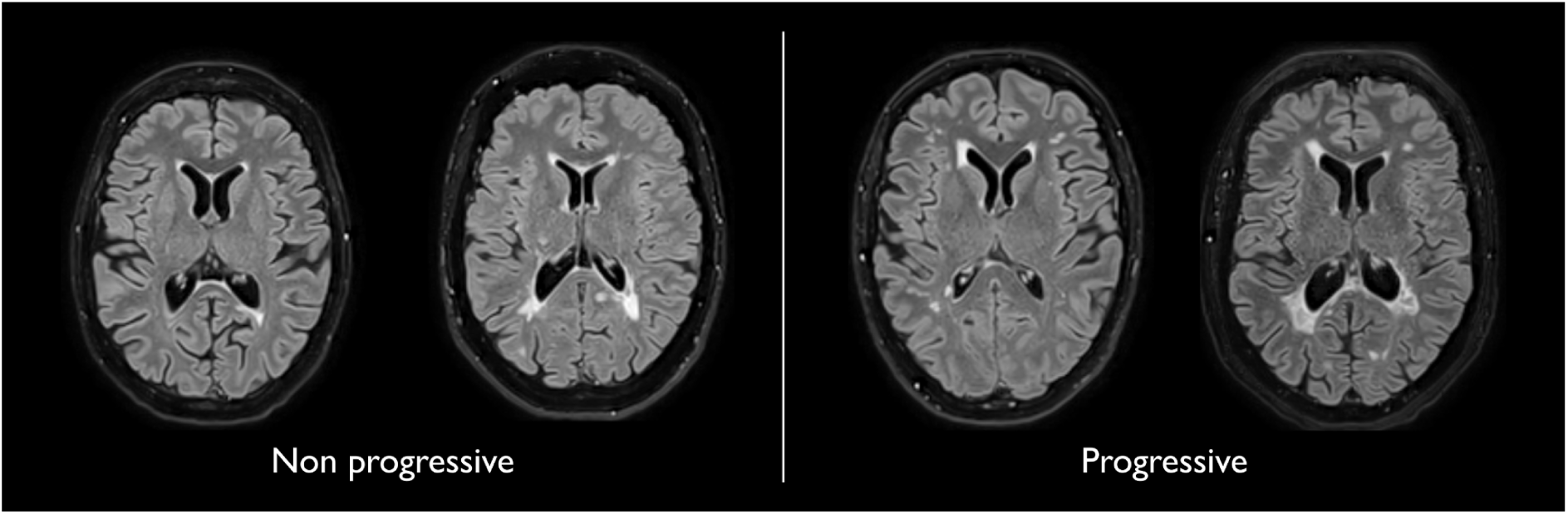
Representative T2-weighted Fluid-attenuated Inversion Recovery (FLAIR) magnetic resonance imaging (MRI) scans of clinically non-progressive (left) and progressive (right) people with multiple sclerosis (PwMS).

The main objective of this study was to predict long-term disability progression in PwMS over a two-year period. Disability progression, derived from changes in the expanded disability status scale (EDSS) (Kurtzke, 1983), corresponds to the worsening of physical capabilities. In clinical practice, EDSS and MRI scans are commonly used to diagnose and assess MS disease course (Wattjes et al., 2021). However, both have their respective limitations. EDSS is prone to inter-rater variability and lacks sensitivity to short-term changes while MRI features, such as the number and volume of white matter lesions (WML) or their gadolinium enhancement, can explain only a fraction of the clinical outcomes in MS (Uitdehaag, 2018). Moreover, these measures fail to track the diffuse pathological changes in the gray matter (GM) and normal-appearing white matter (NAWM) (Davda et al., 2019; Treaba et al., 2019; Wattjes et al., 2021). Therefore, there is an unmet clinical need for more sensitive and specific biomarkers to predict disability progression in PwMS.

An image quantification approach such as radiomics can contribute to potentially overcoming this gap (Pontillo et al., 2021). Radiomics is an automated imaging data quantification approach aimed at extracting high dimensional quantitative features from regions of interest (ROIs) within medical images, such as features related to shape, intensity, texture, etc., to characterise the underlying biology and establish a correlation with clinical and biological endpoints (Gillies et al., 2016; Lambin et al., 2012; Lambin, Leijenaar, Deist, Peerlings, De Jong, et al., 2017; Rogers et al., 2020).

Radiomics has shown promising results in different disease domains such as oncology (Lambin, Zindler, et al., 2017; van Timmeren et al., 2017), Alzheimer’s disease (Feng et al., 2018; Li et al., 2019), and epilepsy (Liu et al., 2018). Similarly, in MS, radiomics features can potentially become clinically relevant non-invasive disease biomarkers for MS disease progression (Lavrova et al., 2021). Some of these features include cortical lesion volume (Calabrese et al., 2010), spinal and brain volume atrophy (Kearney et al., 2015; Storelli et al., 2018), microstructural damage of NAWM (Moll et al., 2011), and the structural changes in the GM (Pontillo et al., 2019).

Given the limitations in EDSS and MRI, training advanced machine learning (ML) techniques with radiomics features holds promise for developing predictive models. By analysing high-dimensional imaging data, ML models can potentially capture and quantify imaging biomarkers associated with MS progression, offering a more robust, objective, and sensitive prediction of long-term disability.

In this study, we used ML to predict long term disability progression in PwMS using real-world data (RWD). Catering to the unmet clinical need to identify a non-invasive quantitative biomarker which is sensitive to disability progression in MS, we hypothesise the following:

- Radiomics based ML models can outperform models relying solely on clinical variables to predict disability progression in PwMS.
- Radiomics features from MRI can predict long-term (2 years) disability progression in PwMS.
- Short-term changes in radiomics features can predict long-term disability progression in PwMS.

## Materials and Methods

### Inclusion Criteria

To ensure the longitudinal tracking of disability progression, inclusion criteria were designed to capture both clinical and imaging data at consistent intervals. This approach was necessary to align MRI scans with corresponding EDSS measurements over time, providing a comprehensive assessment of disease progression. Subjects with a confirmed diagnosis of MS, at least a two-year longitudinal EDSS score trajectory, and at least one baseline and one follow-up MRI scan were included in the study. To achieve this, anchor dates were defined as fixed reference points based on visits (e.g., MRI acquisition dates) and fixed time points (e.g., 6 months and 2 years after the initial visit). Temporal windows were specified around the anchor dates to select follow-up MRI. The three temporal windows were defined as:

- **T0 (initial visit)**: The anchor date for T0 is the MRI acquisition date. The closest EDSS measurement within a 6-month window before or a 3-month window after the T0 anchor date was selected as the baseline score EDSS_T0.
- **T1 (short-term follow-up)**: The anchor date for T1 was set exactly 6 months after the T0 anchor date. The T1 window spanned 3 months before and 3 months after the T1 anchor date. Thus, the MRI session at T1 was selected to calculate short-term changes.
- **T2 (long-term follow-up)**: The anchor date for T2 was set 2 years after the T0 anchor date. The T2 window spanned from 3 months before to 1 year after the T2 anchor date. The closest EDSS measurement to the T2 anchor date was selected as EDSS_T2. No MRI was included in this window, as T2 was based solely on EDSS to assess long-term disability progression.

This structure allowed for consistent alignment of MRI data and EDSS scores at initial visit (T0) and short-term follow-up (T1), while long-term progression (T2) was assessed using EDSS alone. Multiple MRI sessions and their corresponding EDSS scores were included for some subjects, ranging from 2 to 8 MRIs per subject. Each MRI session contributed to the analysis and was treated as a separate observation. Furthermore, MRI sessions from the same subject were grouped during partitioning into training, validation and test sets to avoid potential data leakage. An overview of the temporal windows is illustrated in Figure 2.

**Figure 2:**
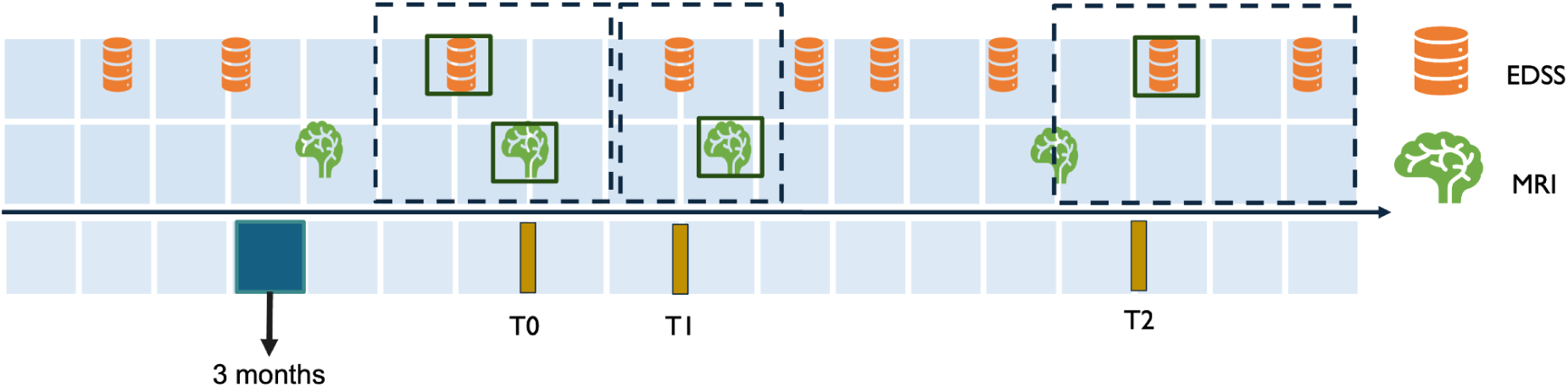
Temporal Alignment of EDSS and MRI Data for Inclusion Criteria. This figure illustrates the temporal windows used to align MRI and EDSS data for assessing disability progression. T0 represents the baseline MRI session, with the closest EDSS measurement selected within a window of 3 months after or 6 months before the MRI. T1 corresponds to the short-term follow-up 6 months after T0, with a 3-month window before and after T1 anchor date for EDSS and MRI selection. T2 represents the long-term follow-up 2 years after T0, with the EDSS measurement selected within a window of 3 months before to 1 year after T2 anchor date.

### Endpoint Definition

The primary endpoint of this study was long-term disability progression, and its definition was adapted from previous work (Kalincik et al., 2017). Long-term disability progression was determined by comparing EDSS scores between T0 and T2. A subject was considered to have worsened if the change in EDSS met the following criteria:

- EDSS_T2 - EDSS_T0 ≥ 1.5 for patients with EDSS_T0 = 0,
- EDSS_T2 - EDSS_T0 ≥ 1.0 for patients with EDSS_T0 ≤ 5.5, or
- EDSS_T2 - EDSS_T0 ≥ 0.5 for patients with EDSS_T0 > 5.5.

### Modelling Pipeline

The pipeline consists of three main stages: image processing, feature processing, and modeling. The image processing stage preprocessed MRI data through reorientation, denoising, bias field correction, super-resolution reconstruction, and segmentation of WML and NAWM. The feature processing stage focused on extracting radiomics features, harmonising them to reduce inter-scanner variability, and removing redundant features. Finally, the modeling stage explored four predictive approaches—clinical, baseline imaging, longitudinal imaging, and combined—by employing feature selection, machine learning models, and validation techniques to evaluate predictive performance. As described later in the section “Data Partitioning”, the datasets were shuffled and split into training, validation, and testing datasets. Feature harmonisation, reduction, selection, model hyperparameters optimisation and model selection was performed exclusively on the training and validation datasets to avoid data leakage.

### Image Processing

#### MRI Data

Our study used pseudonymised longitudinal MRI data collected retrospectively from two medical centres: the Rehabilitation and MS Center of Noorderhart in Pelt, Belgium (DS1) and Zuyderland Medical Center in Sittard, the Netherlands (DS2). The study has been approved by the ethical commission of the University of Hasselt (CME2019/046) and the Medical Ethics Review Committee of Zuyderland and Zuyd University of Applied Sciences (METCZ20200167). No consent to participate was required, given the pseudonymised and retrospective nature of the study. Both DS1 and DS2 have been used for the first time in this study and are private datasets consisting of T2-weighted Fluid-attenuated Inversion Recovery (FLAIR) MRI scans and clinical data, including age, gender, and EDSS scores, collected during routine clinical follow-ups. After applying the inclusion criteria, a total of 149 subjects from DS1 and 98 subjects from DS2 were included in the analysis. The details of the dataset are mentioned in Table 1.

**Table 1:** Datasets summary details.

| **Dataset** | **DS1** | **DS2** | **p-value** |
| --- | --- | --- | --- |
| Participants (n) | 149 | 98 | - |
| Sessions | 630 | 184 | - |
| Age in years at baseline | 42.9 ± 11.7 | 37.4 ± 10.2 | **0.015\*** |
| Female to Male ratio | 3.2:1 | 3.3:1 | 0.983 |
| EDSS at baseline | 2.45 ± 1.56 | 2.87 ± 1.74 | 0.242 |
| Acquisition date range | 2010 - 2017 | 2008 - 2019 | - |
| Worsening (%) | 5.5% | 13% | 0.087 |

The acquisition protocol for T2-weighted FLAIR varied within and across the two datasets. Images in DS1 were acquired using the same scanner (Philips Achieva 1.5T) with three different protocols depending on the date. Between 2010 and 2015, the MRI session included two orthogonal multi-slice T2-W FLAIR acquired with axial and sagittal slice orientations, and with a slice spacing of 6mm (Protocol A). Between 2015 and 2017, three orthogonal images were acquired with a slice spacing of 3mm (Protocol B). In 2017, acquisition sessions included a fast high-resolution 3D T2-W FLAIR with voxel size of 0.98 x 0.98 x 0.6 mm^3 (Protocol C), and most of them also included a structural T1-W MRI (Protocol CsT1). Images in DS2 were acquired in nine different scanners with varying protocols, including low-resolution multi-slice and high-resolution 3D acquisitions, with spacing between slices ranging from 0.8mm to 7mm. Sessions in DS2 were categorised similarly to sessions in DS1, based on the number and resolution of acquired T2-W FLAIR images. Sessions with a 3D high-resolution image were classified as Protocol C, while those with two orthogonal multi-slice low-resolution images were classified as Protocol A. No sessions contained three orthogonal images, and 29 sessions with only one low-resolution image were classified Protocol D. Detailed information about T2-W FLAIR MRI acquisition protocols for both datasets is provided in Appendix A.

#### MRI Pre-processing

The pre-processing of MRI data aimed at harmonising and enhancing the image quality before radiomics feature extraction. First, all MRI images were denoised using adaptive non-local means (Manjón et al., 2010) and N4 bias-field correction (Tustison et al., 2010) was applied to mitigate low-frequency intensity inhomogeneities in MRI images caused by magnetic field distortions. This correction aims at improving the accuracy of segmentation and feature extraction, as it reduces the impact of scanner heterogeneity (Tustison et al., 2010).

For protocols with low-resolution images (A, B, and D), we applied “perceptual super-resolution in multiple sclerosis” (PRETTIER) (Giraldo et al., 2024), a super-resolution approach designed to enhance the through-plane resolution of multi-slice structural MRIs containing MS lesions. Since protocols A and B have multiple low-resolution FLAIR images per session, we applied PRETTIER to each image and aligned and combined the outputs following an iterative approach. This technique improves spatial resolution, which is important for downstream radiomics analysis and segmentation tasks. Reconstruction was not performed on protocol C since it had high-resolution FLAIR.

Next, we applied the Sequence Adaptive Multimodal SEGmentation (SAMSEG) method for whole-brain segmentation on all FLAIR protocols across DS1 and DS2 (Cerri et al., 2021). SAMSEG is a previously validated segmentation tool designed to segment 41 anatomical brain structures (See Appendix B) from MRI and is fully adaptive to different MRI contrasts and scanners, making it particularly suitable for multi-center datasets like ours. SAMSEG was used to segment, among others, the normal-appearing white matter (NAWM), gray matter (GM), thalamus and cerebrospinal fluid (CSF).

Lesions were segmented using the lesion prediction algorithm (Schmidt, 2017) as implemented in LST toolbox version 1.2.3 (www.statistical-modelling.de/lst.html) for SPM8 (http://www.fil.ion.ucl.ac.uk/spm). This algorithm uses a pre-trained logistic regression model to generate lesion probability estimates at each voxel. These lesion probability estimates were thresholded at 0.1 to create white matter lesions (WML) masks. The flow chart of the entire pipeline is shown in Figure 3.

**Figure 3.**
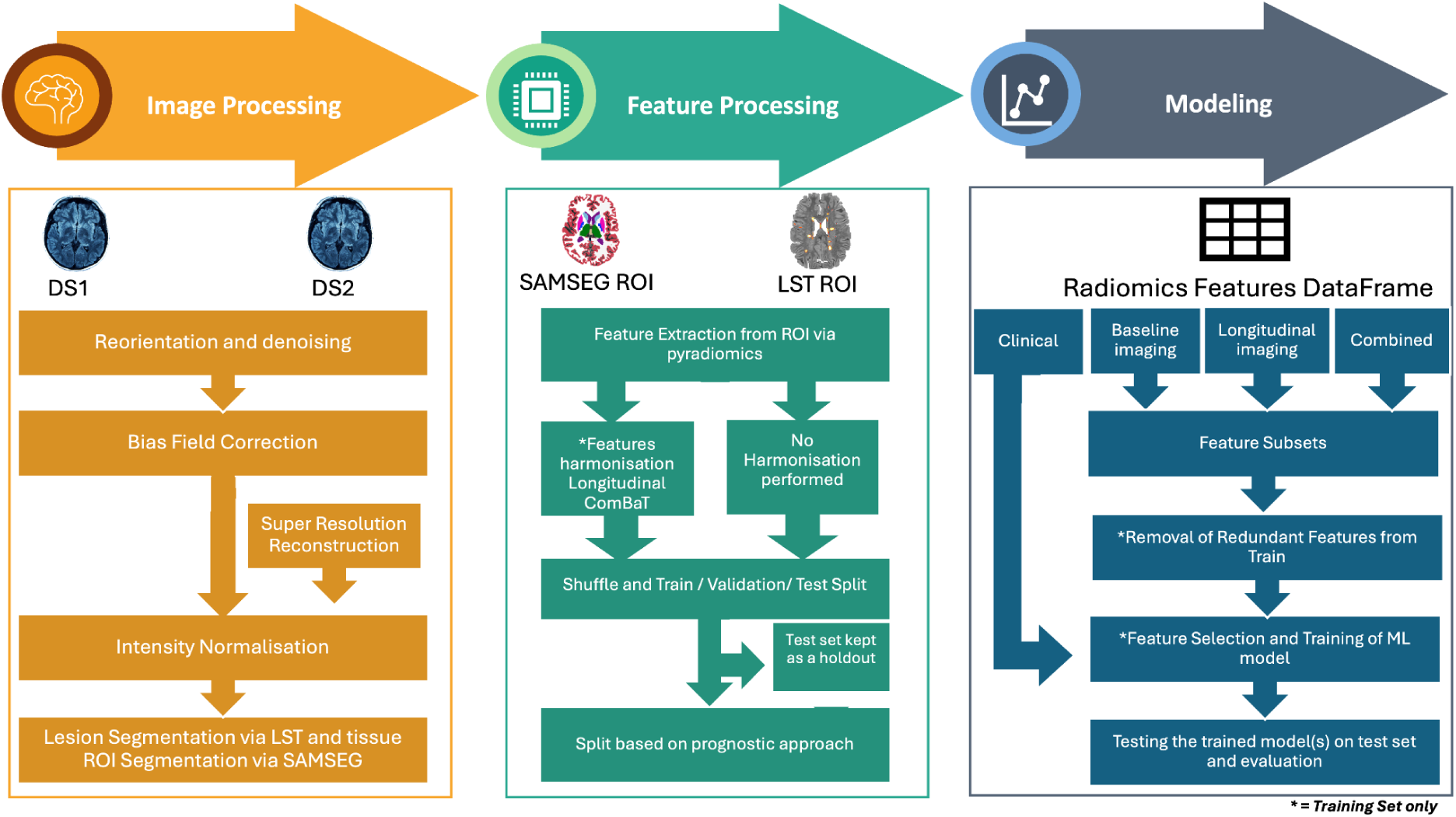
Overview of the Methodology for MRI-Based Disability Progression Prediction in Multiple Sclerosis. The pipeline consists of three main stages: Image Processing, Feature Processing, and Modeling. Image processing involves image pre-processing steps such as reorientation and denoising, bias field correction, intensity normalisation, and lesion/tissue segmentation using LST and SAMSEG. Super-resolution reconstruction was applied to low-resolution images. Feature processing includes the extraction of radiomics features from regions of interest (ROI) and harmonisation using longitudinal ComBat to address inter-scanner variability. Features were then divided into harmonised and non-harmonised datasets. The feature sets extracted from DS1 and DS2 were then shuffled and divided into training, validation, and test sets. Modeling stage includes the division of dataset according to four prognostic approaches Clinical, Baseline imaging, Longitudinal imaging and Combined. This was followed by further subdividing each approach into feature subsets and subsequent removal of feature reduction, feature selection and training of machine learning model, and finally evaluating the best model on the test set.

### Feature Processing

Besides having masks as the outcome of segmentation, anatomical and lesion volumes were also obtained using SAMSEG, and LST. These volumes were subsequently normalised by intracranial volume to ensure comparability across subjects. Per-image adaptive histogram matching (Pizer et al., 1987) was then performed to normalise the intensity distributions of all the skull-stripped FLAIR images, ensuring consistency in intensity values across images.

For this study, high dimensional radiomics features were extracted from two regions of interest (ROI): the NAWM and the WML. The WML mask was subtracted from the segmented WM mask to generate the NAWM mask. These binary masks, along with the intensity-normalised FLAIR images, were used to compute the corresponding radiomics features for further analysis.

#### Feature Extraction

Radiomics features from the ROIs were extracted using PyRadiomics 2.20 (van Griethuysen et al., 2017) with python 3.7.1. The extracted radiomics features comprised six classes, including shape (Lorensen & Cline, 1987), first-order statistics (FO), gray-level co-occurrence Matrix (GLCM) (Haralick et al., 1973), gray-level run length matrix (GLRLM) (Galloway, 1975), gray-level size zone matrix (GLSZM) (Thibault et al., 2013) and gray-level dependence matrix (GLDM) (Sun & Wee, 1983). Gray-level features were calculated by discretising the images with 50 bins, which is in line with the recommendations by the Image Biomarker Standardization Initiative (IBSI) (Zwanenburg et al., 2018) and in the documentation of PyRadiomics (van Griethuysen et al., 2017). The details of the number of features extracted per class is available in Appendix K.

#### Feature Harmonisation

Given the diversity of MRI acquisition protocols and the longitudinal heterogeneity of DS1 and DS2, harmonisation of the radiomics features between different protocols was performed using longitudinal ComBat (Beer et al., 2020). This technique was applied to improve comparability of the features across the datasets by minimising inter- and intra-site variability, as well as temporal variations in MRI which can stem from the acquisition protocol. The principal component analysis (PCA) visualisation of the features from DS1 and DS2 before and after harmonisation is illustrated in Appendix H. Harmonisation coefficients were calculated in the training dataset only, and later applied to the testing and validation datasets.

To ensure a comprehensive evaluation, all subsequent steps were conducted separately to both harmonised and non harmonised datasets.

#### Dataset Partitioning

The DS1 and DS2 were shuffled and were then split into training (60%), validation (20%) and test (20%) sets. The splitting was performed using stratified shuffled split using scikit-learn (Pedregosa et al., 2011). This ensured two things: 1) the distribution of subjects with worsening disability progression was stratified across the sets as evenly as possible, and (2) sessions of the same subject were kept within the same dataset, preventing data leakage.

#### Prognostic Approaches

For this study, four distinct prognostic approaches were defined. First, we included a “clinical” approach, focusing solely on routinely available clinical variables such as gender, clinical age (in years) and EDSS at T0. The reason for analysing this approach separately was to evaluate the predictive value of clinical features alone, independent of radiomics features, and served as a baseline for comparison with radiomics based approaches.

Since our study also aimed to capture the predictive capability of both baseline features and short-term feature changes in MRI data for disability progression, we further defined three distinct radiomics-based prognostic approaches. For extracting radiomics features, we used all sessions for each subject with the disability progression label corresponding to that session, labelling this as the “baseline imaging” approach. The second approach, “longitudinal imaging”, focused on short-term changes in all radiomics features relative to the baseline features. This is calculated by dividing the difference between T1 and T0 and dividing it by T0 (Heidt et al., 2024). In this case, the disability progression label corresponded to the one assigned at T0. Lastly, to assess whether combining baseline imaging and longitudinal imaging could improve predictive power, we integrated features used in longitudinal imaging and baseline imaging approaches, referring to this as the “combined” approach, with the disability progression label taken at T0.

### Modeling

#### Feature Subsets

To evaluate further whether radiomics features alone or with clinical data are able to predict to disability progression, we divided the features in baseline imaging, longitudinal imaging and combined prognostic approaches into the following subsets:

- **Radiomics volume features**: Regional and lesion volumes derived from SAMSEG, and LST (normalised by intracranial volume).
- **Radiomics features without volumes**: Features extracted exclusively using PyRadiomics (e.g., shape, FO, GLCM, GLRLM, GLSZM, and GLDM).
- **Radiomics features**: A combination of both the selected radiomics volume features and radiomics features without volumes.
- **Radiomics and clinical features**: A combination of the selected radiomics features with clinical data, including gender (female), clinical age (in years) and EDSS at T0. This was done to test whether radiomics features combined with clinical features enhance predictive power of the model.

#### Feature Selection

Feature reduction and selection was not applied for clinical only dataset, however, for the radiomics-based prognostic approaches, feature reduction was performed separately on the training set for both the harmonised and non harmonised data analysis pipelines. This includes the four feature subsets and the aim was to eliminate redundant and non-informative features. Initially, all features were normalised using StandardScalar from scikit-learn (Pedregosa et al., 2011), and then pairwise Spearman correlation was computed for the entire feature set. Features that exhibited a Spearman correlation coefficient greater than 0.9 were considered highly correlated. From each pair of intercorrelated features, the one with the higher average Spearman correlation across all other features was flagged for removal. To ensure stability in the selection process, a bootstrapping approach was used: in each iteration, stratified subsamples were generated based on the outcome, and the intercorrelated features were recalculated. Features that appeared as candidates for removal in 50% or more of the bootstrap iterations were discarded from the final dataset, resulting in a robust set of non-intercorrelated features for further analysis.

Recursive Feature Elimination with 20-fold cross-validation (RFECV) was applied on the non-intercorrelated features across each approach and feature subset. Given the imbalance in the training dataset, with only 6.9% subjects showing worsening disability, we used a Balanced Random Forest Classifier (BRFC) as an estimator within RFECV to account for this imbalance. Stratified Shuffle Split cross-validation was applied to maintain class proportion across folds, and precision score was chosen as the evaluation metric to prioritise features that improve the precision of disability progression prediction.

#### Selection of Classification Model

We used three ML models, Elastic Net (logistic regression), BRFC, and Light Gradient-Boosting Machine (LGBM), to evaluate each prognostic approach and the feature subsets it entails. In an attempt to make the models robust, we employed Optuna to optimise the hyperparameters of each model. Optuna is a framework for efficient hyperparameter tuning using Bayesian optimisation (Akiba et al., 2019). Moreover a 20-fold cross-validation alongside stratified shuffle split was implemented during model hyperparameter tuning as well. This approach ensured stability by iteratively training and testing models across different subsets of the training data, helping to enhance the generalisability of the predictions. The model that had the best area under the precision-recall curve (PR AUC) on the validation set, per feature subset, was subsequently tested on the test set to evaluate the generalisability of the feature subset per approach. The PR AUC curve was used to select the best-performing models on the validation set, as it emphasises the trade-off between precision (positive predictive value) and recall (sensitivity), making it especially suited for imbalanced datasets, like ours, where worsening cases are sparse. In addition to the PR AUC curve, the area under the receiver operating characteristic curve (ROC AUC) was also used to give an insight into the discriminative capabilities of the models. For PR AUC, a value significantly above the prevalence of the positive class indicates meaningful performance. Whereas for ROC AUC, a value of 0.5 indicates random performance and higher values reflect better discrimination (Çorbacıoğlu & Aksel, 2023).

To get an actual picture of the sensitivity and specificity of the models, we used Youden’s index (J) to calculate the optimal threshold for binary classification (Youden, 1950). Furthermore, to validate whether the results generated by our models are not a result of random chance, we also conducted permutation analysis by shuffling the outcome variable, i.e. the disability progression, and re-ran model training. The permutation was performed 50 times, and the models were then subsequently evaluated.

To interpret the predictions of the ML models, SHapely Additive exPlanations (SHAP) (Lundberg et al., 2019) were employed. SHAP values were computed for the features of selected models, followed by the generation of summary plots to visualise feature importance and their impact on predictions. The plots ranked the features by their mean absolute SHAP values and their respective effect on the likelihood of disability progression.

Moreover, the Radiomics Quality Score (RQS) framework was followed to ensure methodological rigour and adherence to radiomics standards (Lambin, Leijenaar, Deist, Peerlings, de Jong, et al., 2017). The CLEAR (Checklist for Evaluating the Reporting of AI in Radiology) checklist was also used to evaluate the transparency and reproducibility of the ML pipeline, ensuring clarity and alignment with best practices for AI reporting (Kocak et al., 2023).

## Results

### Cohort Characteristics

By combining and shuffling DS1 and DS2, a total of 247 PwMS were included in this study. The 247 participants were divided, using stratified shuffled split, into training (n=148), validation (n=49), and test (n=50) sets. Subjects in the training set had an average age of 41.32 years (±12.13), while those in the validation and test sets had averages of 44.79 years (±10.92) and 38.52 years (±10.25), respectively. The female-to-male ratio remained almost consistent across sets at approximately 3.3:1. Worsening disability progression was observed in 6.9% of the training set, 9.8% of the validation set, and 6.0% of the test set. The summary of cohort characteristics for shuffled datasets has been outlined in Table 2.

**Table 2:** Characteristics of the training, validation and test datasets.

| Dataset | Training Set | Validation Set | Test Set | p-value |
| --- | --- | --- | --- | --- |
| Participants | 148 | 49 | 50 | - |
| Sessions | 470 | 166 | 178 | - |
| Age in years at baseline | 41.32 ± 12.13 | 44.79 ± 10.92 | 38.52 ± 10.25 | **Training vs Validation - 0.04\***<br>**Validation vs Test - 0.01\***<br>Training vs Test - 0.37 |
| Female to Male ratio | 3.2:1 | 3.3:1 | 3.3:1 | Training vs Validation - 0.98<br>Validation vs Test - 0.99<br>Training vs Test - 0.99 |
| EDSS at baseline | 2.55 ± 1.56 | 2.92 ± 1.45 | 2.17 ± 1.50 | Training vs Validation - 0.11<br>**Validation vs Test - 0.04\***<br>Training vs Test - 0.08 |
| Percentage Split (of total) | 60 % | 20% | 20% | - |
| Acquisition date range | 2008 - 2019 | 2010 - 2019 | 2010 - 2019 | - |
| Worsening (%) | 6.9% | 9.8% | 6.0% | Training vs Validation - 0.67<br>Validation vs Test - 0.58<br>Training vs Test - 0.84 |

The longitudinal imaging approach resulted in a dataset reduction of unique participants and sessions because it was constructed by calculating feature differences between T0 and T1 sessions. The characteristics of delta datasets are summarised in Table 3.

**Table 3:**
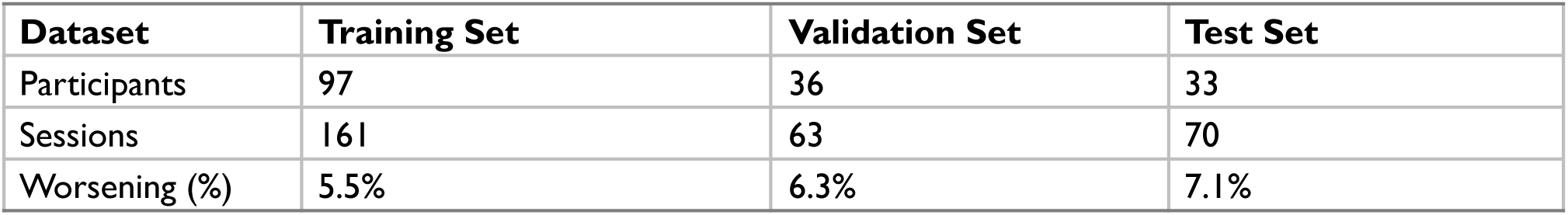
Characteristics of longitudinal imaging approach training, validation and test datasets.

| Dataset | Training Set | Validation Set | Test Set |
| --- | --- | --- | --- |
| Participants | 97 | 36 | 33 |
| Sessions | 161 | 63 | 70 |
| Worsening (%) | 5.5% | 6.3% | 7.1% |

### Feature Selection

Tissue segmentation using SAMSEG produced 41 anatomical features (volumes only) from different brain regions. Out of 41, two features: “unknown volumes” and “fifth ventricle volume” were dropped due to its negligible size in the MRI. The remaining 39 features, along with 2 lesion-specific features from LST, the number of lesions and lesion volume, constituted the radiomics volume feature subset. Instances where LST failed to provide the lesion volume feature, the SAMSEG derived WML volume was used as a substitute. Additionally, high-dimensional radiomics features extracted using PyRadiomics, yielded a total of 200 features for the radiomics features without volumes subset.

Subsequently, per prognostic approach, intercorrelated features were dropped, the details of which are summarised in Appendix C. All the unique retained non-intercorrelated features underwent RFECV to get the optimum number of features for downstream ML analysis. The feature subsets selected by RFECV for the best performing models per approach are shown in Table 3.

**Table 3a.**
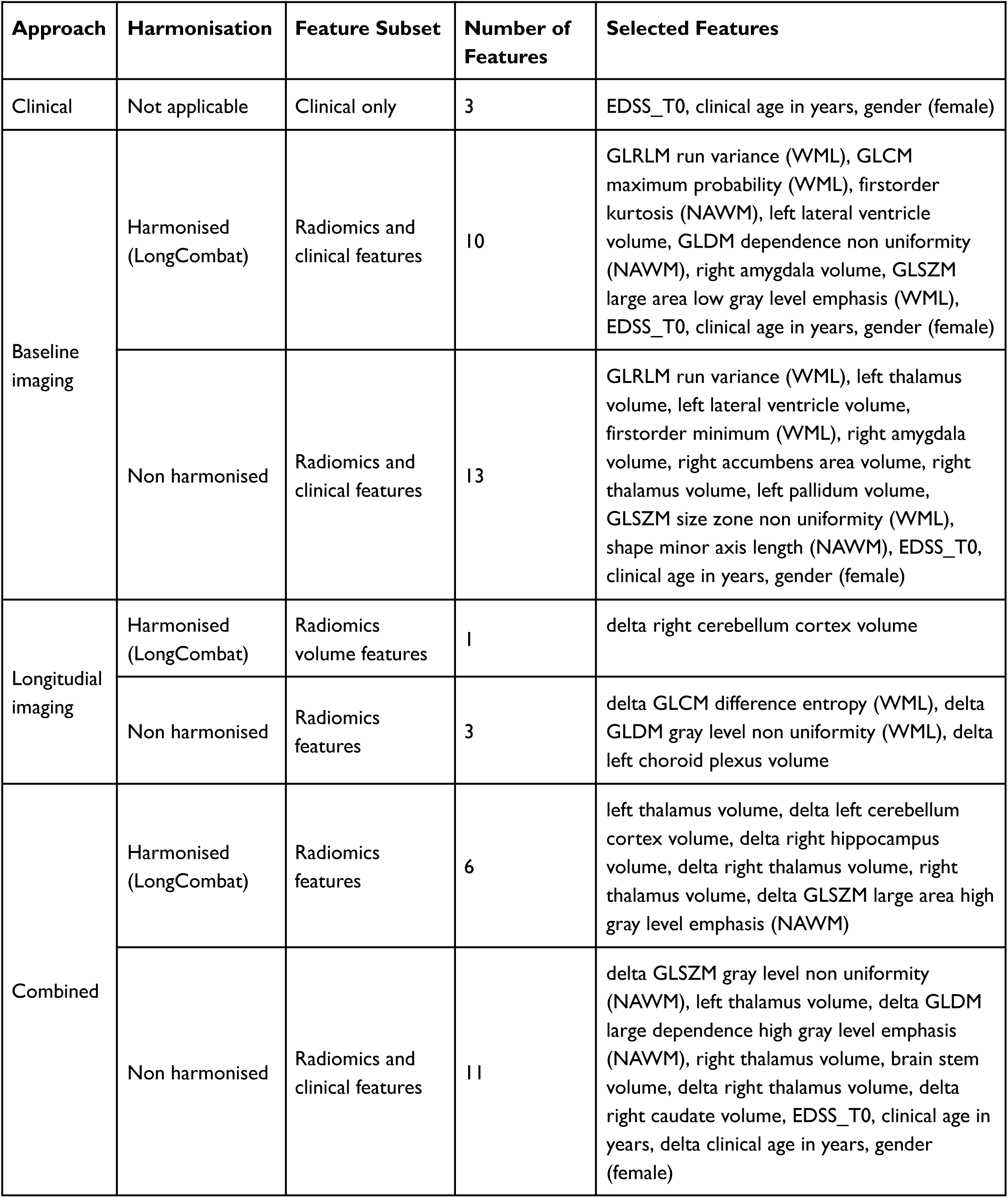
Selected features from the best-performing models for disability progression prediction.

### Machine Learning Models Performance

The results of the best ML model per prognostic approach is summarised in Table 4. As shown in figures 4 and 5, for the clinical approach, LGBM performed the best by achieving a validation PR AUC of 0.12 and a validation ROC AUC of 0.57. On the test set, it attained a PR AUC of 0.08 and a ROC AUC of 0.6.

**Figure 4.**
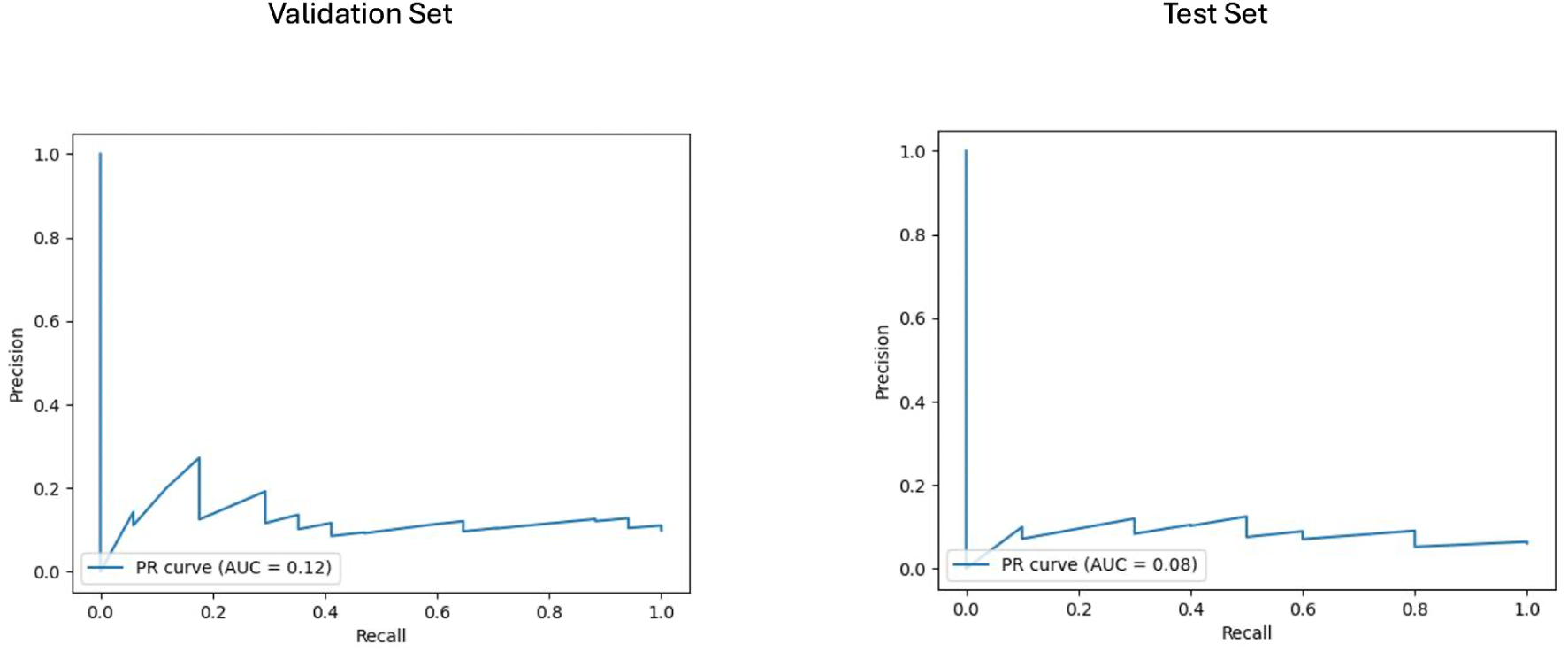
Area under the precision-recall curve (PR AUC) or model performance on clinical validation and test sets. The PR curves illustrate the performance of the best-performing model LGBM model trained on clincial features for predicting disability progression. Validation PR AUC was 0.12, while the test set PR AUC was 0.08.

**Figure 5.**
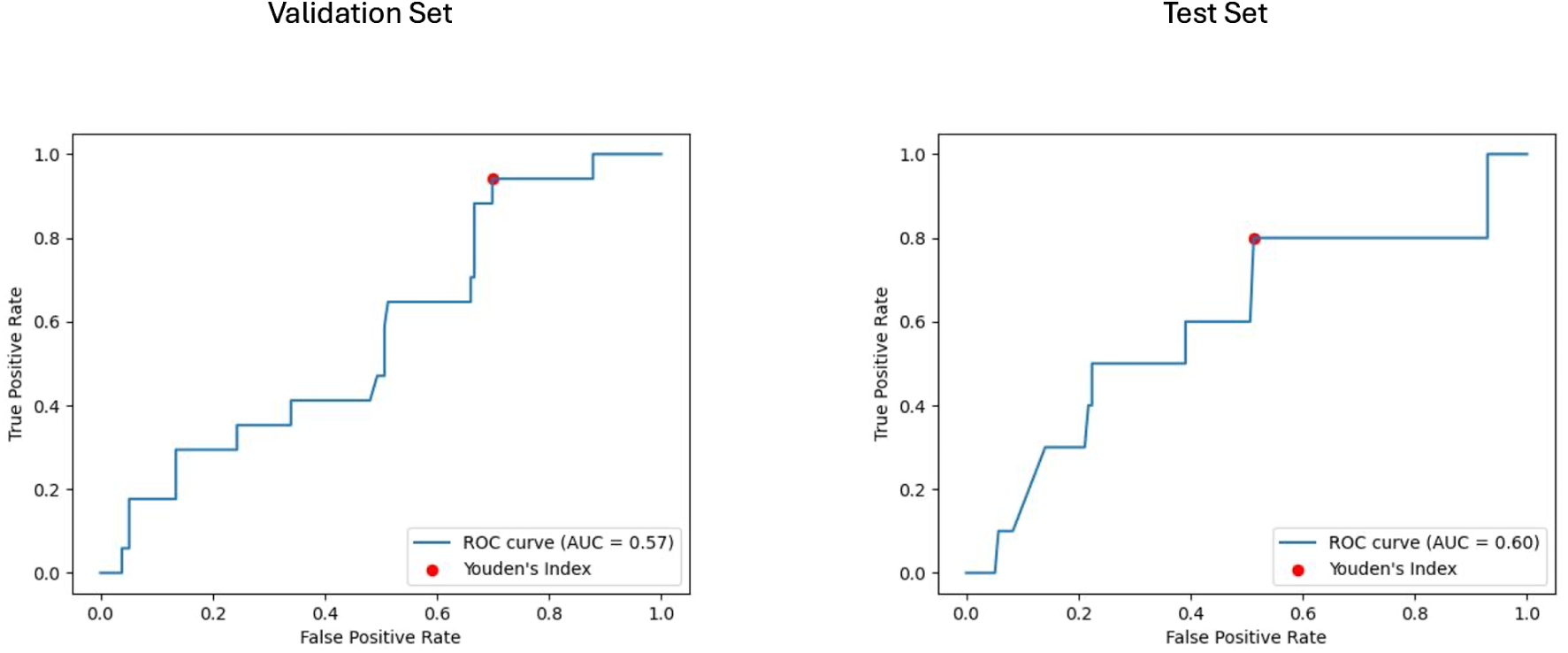
Receiver operating characteristic curve with area under the curve (ROC AUC) for model performance on clinical validation and test set. The ROC curves display the discriminative ability of the best-performing LGBM model trained on clinical features for predicting disability progression. The ROC AUC for the validation set was 0.57, with a test set ROC AUC of 0.60. The red dots indicate the optimal thresholds determined using Youden’s index, balancing sensitivity and specificity.

**Table 4:**
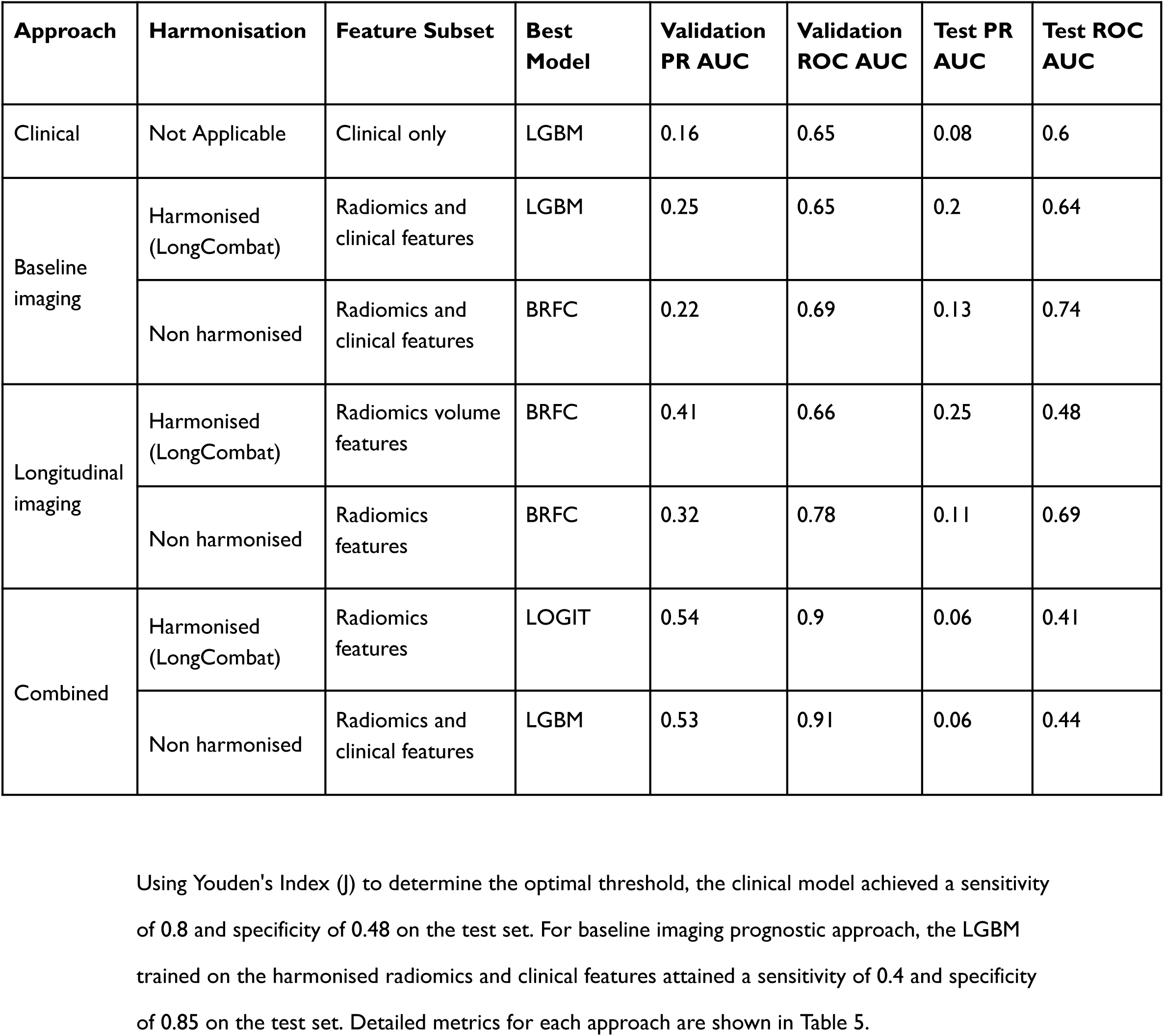
Performance metrics of the best-performing models across approaches, harmonisation strategies, and feature subsets.

| Approach | Harmonisation | Feature Subset | Best Model | Validation PR AUC | Validation ROC AUC | Test PR AUC | Test ROC AUC |
| --- | --- | --- | --- | --- | --- | --- | --- |
| Clinical | Not Applicable | Clinical only | LGBM | 0.16 | 0.65 | 0.08 | 0.6 |
| Baseline imaging | Harmonised (LongCombat) | Radiomics and clinical features | LGBM | 0.25 | 0.65 | 0.2 | 0.64 |
|  | Non harmonised | Radiomics and clinical features | BRFC | 0.22 | 0.69 | 0.13 | 0.74 |
| Longitudinal imaging | Harmonised (LongCombat) | Radiomics volume features | BRFC | 0.41 | 0.66 | 0.25 | 0.48 |
|  | Non harmonised | Radiomics features | BRFC | 0.32 | 0.78 | 0.11 | 0.69 |
| Combined | Harmonised (LongCombat) | Radiomics features | LOGIT | 0.54 | 0.9 | 0.06 | 0.41 |
|  | Non harmonised | Radiomics and clinical features | LGBM | 0.53 | 0.91 | 0.06 | 0.44 |

For the baseline imaging approach LGBM performed best in the radiomics and clinical features subset. As shown in Figure 6 and Figure 7, LGBM achieved a validation PR AUC of 0.28 and a validation ROC AUC of 0.73. On the test set, it attained a PR AUC of 0.20 and an ROC AUC of 0.64. While the non hamornised baseline models also generalised well on the test set, it however, did not achieve better results compared to the baseline harmonised model (See Appendix D).

**Figure 6.**
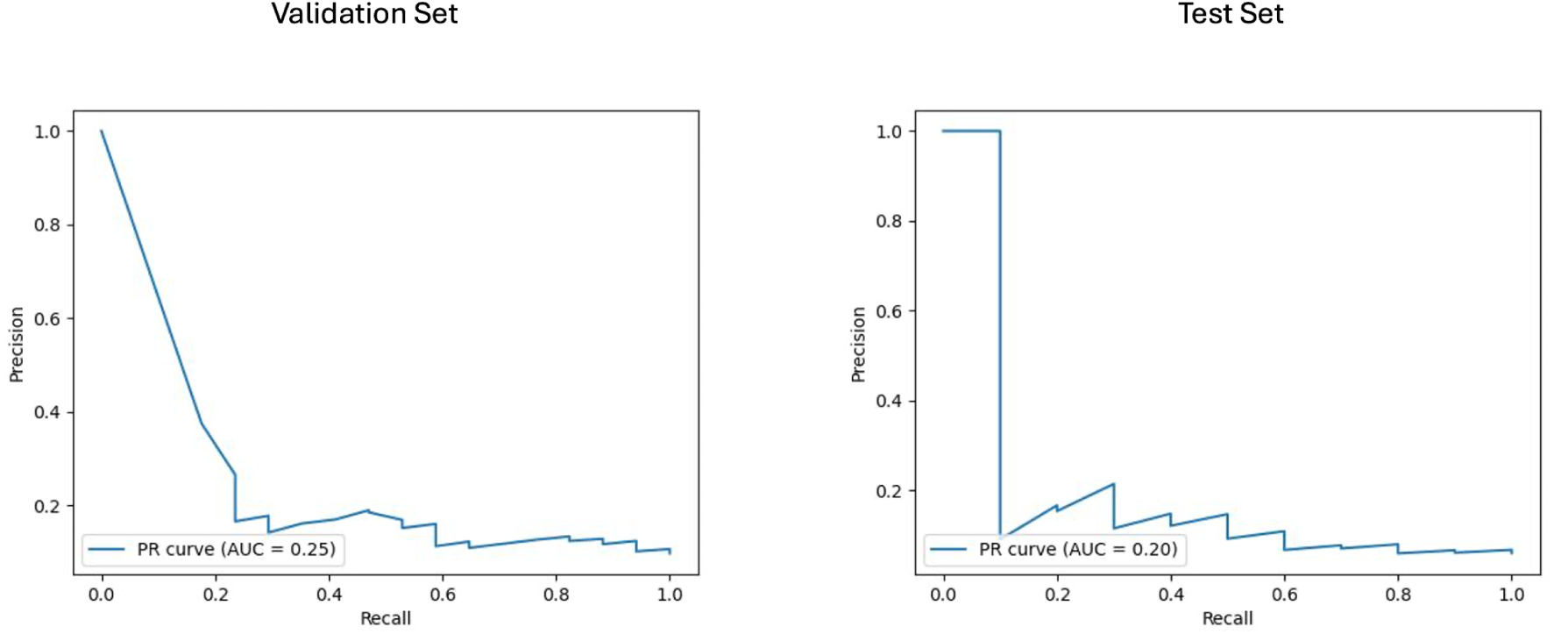
Area under the precision-recall curve (PR AUC) for model performance on baseline imaging validation and test sets with harmonised radiomics and clinical feature subset. The PR curves illustrate the performance of the best-performing model LGBM trained on baseline imaging harmonised radiomics and clinical feature subset for predicting disability progression. Validation PR AUC was 0.25, while the test set PR AUC was 0.20.

**Figure 7.**
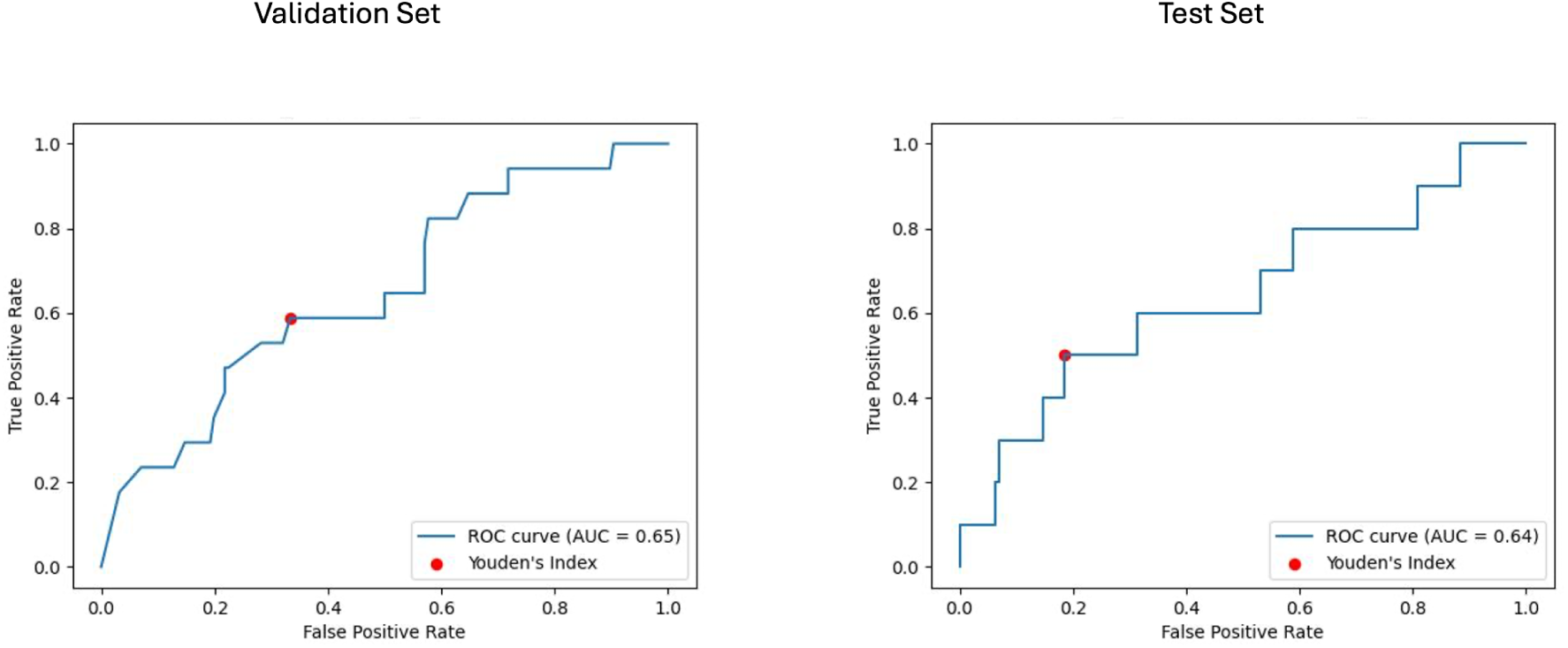
Receiver operating characteristic curve with area under the curve (ROC AUC) for model performance on baseline imaging validation and test sets with harmonised radiomics and clinical feature subset. The ROC curves display the discriminative ability of the best-performing LGBM model trained on baseline imaging harmonised radiomics and clinical feature subset. The ROC AUC for the validation set was 0.65, with a test set ROC AUC of 0.64. The red dots indicate the optimal thresholds determined using Youden’s index, balancing sensitivity and specificity.

For the longitudinal imaging prognostic approach, the BRFC model trained on non-harmonised radiomics features achieved the best results compared to the harmonised approach, with a validation PR AUC of 0.32 and ROC AUC of 0.78, while on the test set, it achieved a PR AUC of 0.11 and an ROC AUC of 0.69 (See figures 8 and 9). The longitudinal imaging harmonised models and the combined models, both harmonised and non harmonised, did not generalise well on the test set (see Table 4).

**Figure 8.**
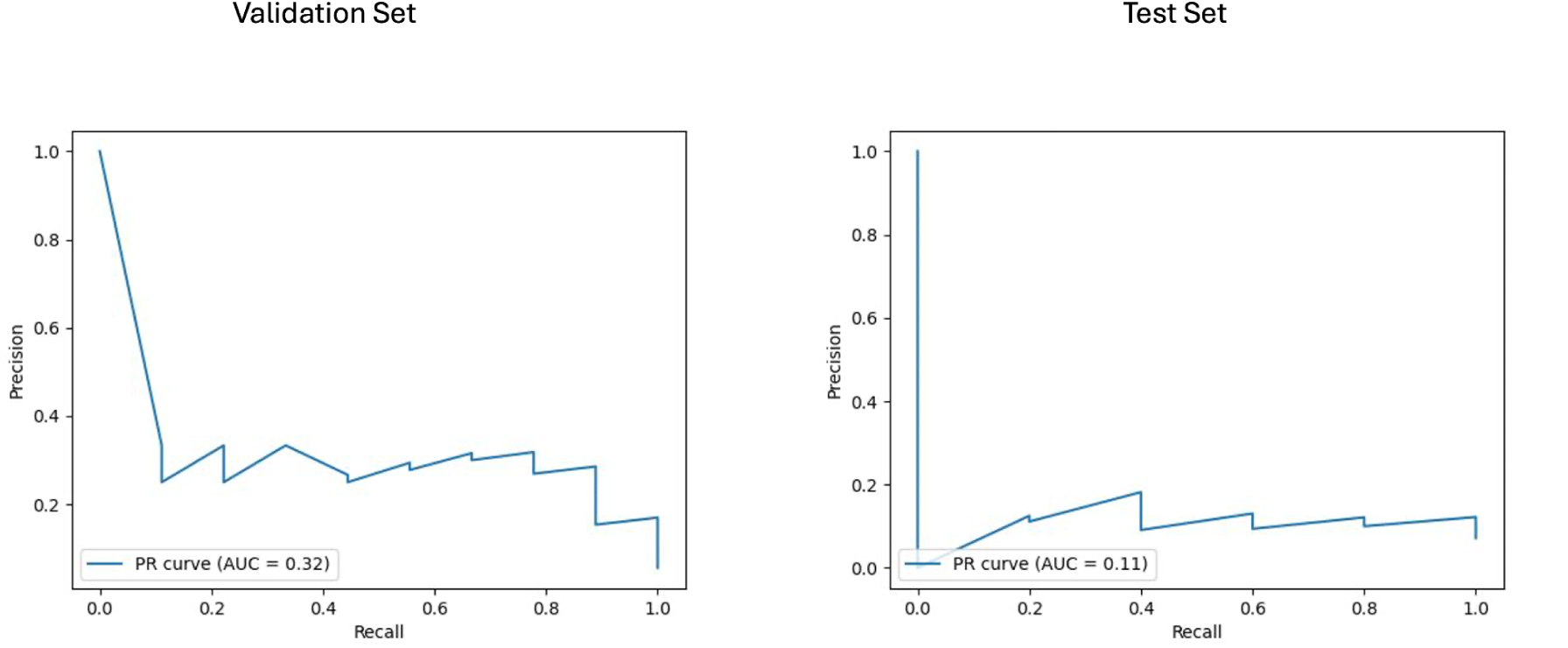
Area under the precision-recall curve (PR AUC) for model performance on longitudinal imaging validation and test sets with non-harmonised radiomics feature subset. The PR curves depict the performance of the best-performing model, BRFC, for longitudinal imaging non-harmonised radiomics feature subsets. The validation PR AUC is 0.32 and test PR AUC is 0.11.

**Figure 9.**
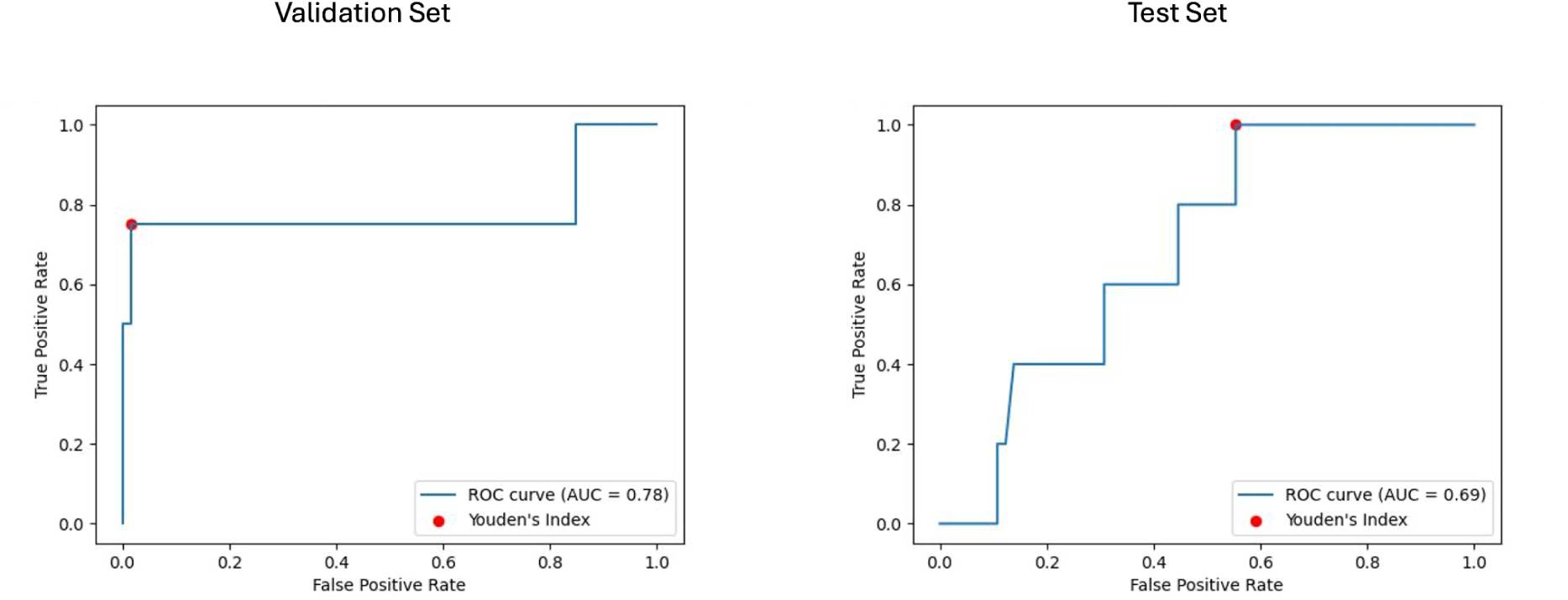
Receiver operating characteristic curve with area under the curve (ROC AUC) for model performance on longitudinal imaging validation and test sets with non-harmonised radiomics feature subset. The ROC curves display the discriminative ability of the best-performing BRFC model for longitudinal imaging non-harmonised radiomics feature subsets. The validation ROC AUC reached 0.78, while the test set ROC AUC was 0.69. The red dots indicate the optimal thresholds determined using Youden’s index, balancing sensitivity and specificity.

Using Youden’s Index (J) to determine the optimal threshold, the clinical model achieved a sensitivity of 0.8 and specificity of 0.48 on the test set. For baseline imaging prognostic approach, the LGBM trained on the harmonised radiomics and clinical features attained a sensitivity of 0.4 and specificity of 0.85 on the test set. Detailed metrics for each approach are shown in Table 5.

**Table 5.** Sensitivity, specificity, precision and recall metrics for the best-performing models determined by Youden’s index.

| Approach | Harmonisation | Feature Subset | Best Model | Youdens Index | Sensitivity | Specificity | Precision | Recall |
| --- | --- | --- | --- | --- | --- | --- | --- | --- |
| Clinical | Not Applicable | Clinical only | LGBM | 0.03 | 0.8 | 0.48 | 0.09 | 0.8 |
| Baseline imaging | Harmonised (LongCombat) | Radiomics and clinical features | LGBM | 0.0086 | 0.5 | 0.81 | 0.15 | 0.5 |
| Longitudinal imaging | Non harmonised | Radiomics features | BRFC | 0.23 | 1.0 | 0.45 | 0.12 | 1.0 |

### SHAP based feature analysis

The SHAP analysis identified the most influential features contributing to the prediction of disability progression across the best-performing models in the baseline imaging and longitudinal imaging prognostic approaches. For the baseline imaging prognostic approach with harmonised radiomics and clinical features, as shown in figure 10, the SHAP analysis revealed GLCM maximum probability (WML), left lateral ventricle volume, and GLDM dependence non-uniformity (NAWM) as the top three features influencing predictions. Features like gender (female) had a lower impact on the model outcome.

**Figure 10.**
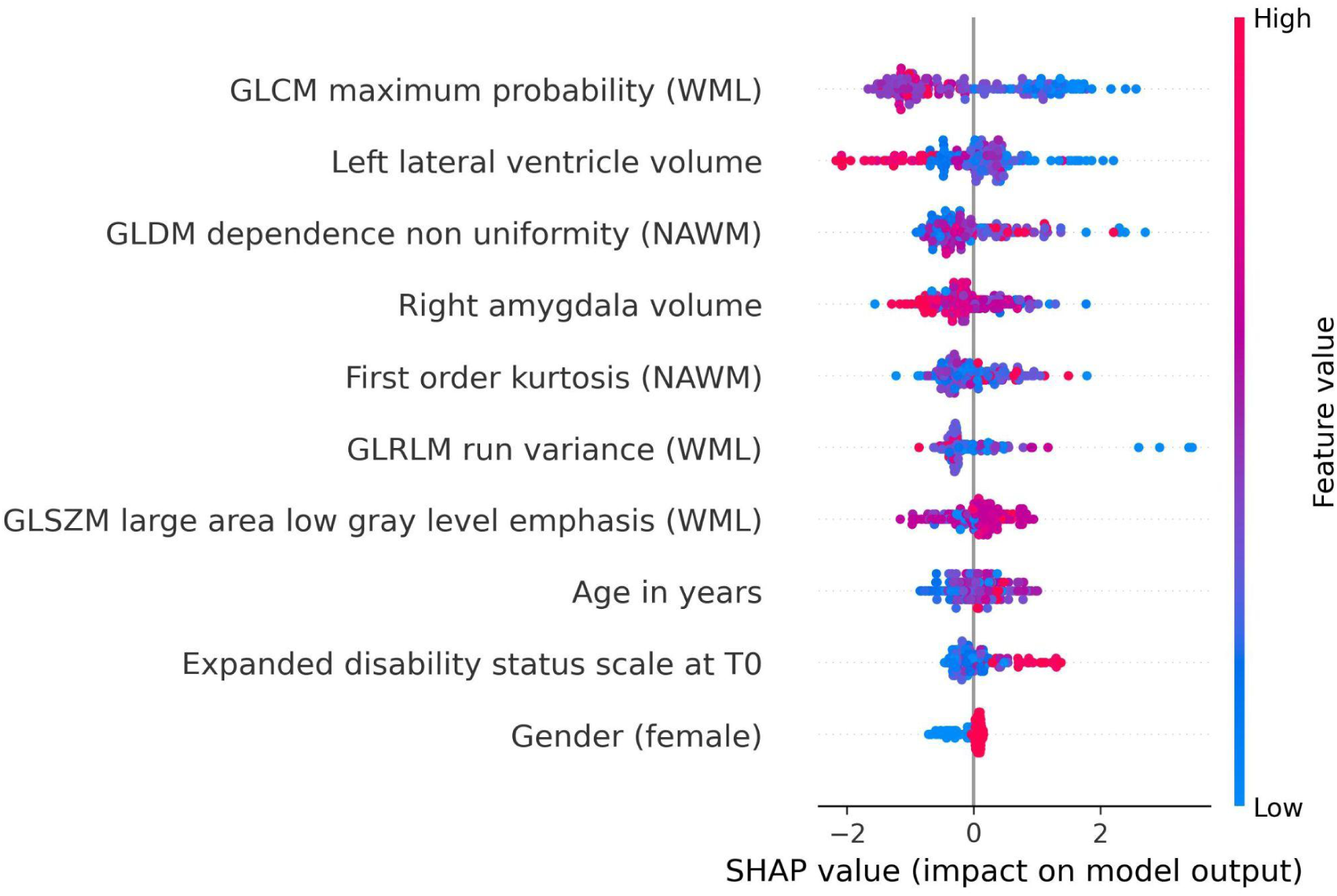
SHAP summary plot for model trained using baseline imaging prognostic approach with harmonised radiomics and clinical feature subset. The SHAP analysis highlights the top contributing features influencing the prediction of disability progression. Key features include GLCM maximum probability (WML), left lateral ventricle volume, and GLDM dependence non uniformity (NAWM), reflecting the importance of textural and anatomical characteristics in predicting progression. Features like gender (female) and age at baseline had relatively lower contributions to the model’s predictions.

In the SHAP summary plot (figure 10 & 11), features are ranked by their mean absolute SHAP value, which quantifies their overall importance in the model. The higher the mean absolute SHAP value, the greater the feature’s contribution to predictions across all subjects. The color coding in the plot represents the value of the feature for each individual subject: red points correspond to higher feature values, while blue points indicate lower feature values. For example, higher GLCM maximum probability (red points) was associated with a lower likelihood of disability progression, reflecting its inverse relationship with the outcome.

**Figure 11.**
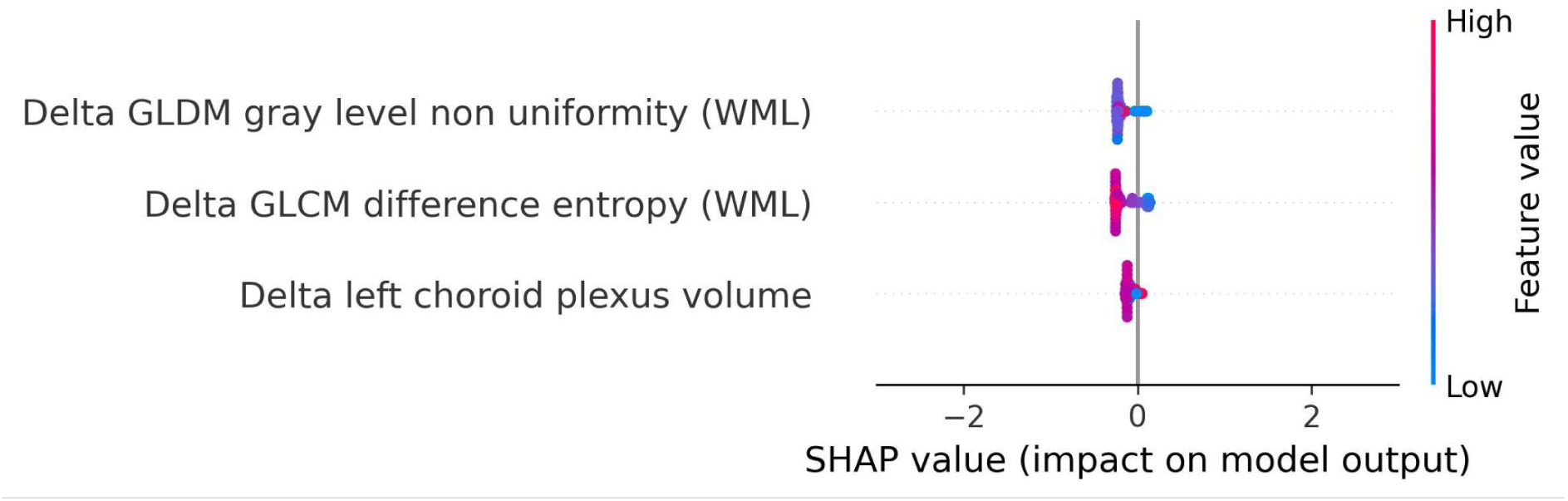
SHAP summary plot for model trained using longitudinal imaging prognostic approach with non harmonised radiomics feature subset. The SHAP analysis for the delta non-harmonised model identifies delta GLDM gray level non-uniformity (WML) and delta GLCM difference entropy (WML) as the most predictive features for disability progression. The delta left choroid plexus volume exhibited a lower contribution, underscoring the relative importance of dynamic changes in lesion structure over time.

For the longitudinal imaging approach with non-harmonised radiomics feature subset, the delta GLDM gray level non-uniformity (WML) and delta GLCM difference entropy (WML) had a higher predictive capability for disability progression, whereas the delta left choroid plexus volume had a relatively lower contribution to the model’s outcome (see figure 11).

For details on the specific model parameters and settings we refer to Appendix I, whereas the details of CLEAR checklist and RQS are provided in Appendix E and Appendix F respectively.

The results of the permutation testing conducted by shuffling the outcome variable and re-evaluating the models are presented in Appendix J. The findings show that the performance of the permuted models in all the prognostic approaches was worse compared to the original models.

## Discussion

In this study, we explored the potential of FLAIR MRI-based radiomics and ML techniques on multicentric data to predict disability progression in people with multiple sclerosis. We deployed three ML models, namely LOGIT, BRFC and LGBM, across four different prognostic approaches, i.e., clinical, baseline imaging, longitudinal imaging and combined. Except for the clinical approach, the imaging and combined prognostic approaches further consist of harmonised and non-harmonised feature subsets comprising radiomics volume features, radiomics features without volumes, radiomics features, as well as radiomics and clinical features subset.

Addressing our first research question, whether radiomics based models can outperform models relying solely on clinical variables, we found that LGBM model trained of harmonised radiomics and clinical features generalised the best on test set, achieving a PR AUC of 0.2 and ROC AUC of 0.64 on the test set. As shown in Appendix D, the combination of radiomics and clinical features outperformed both the clinical-only and radiomics-only prognostic approaches, demonstrating the added value of integrating advanced imaging biomarkers with routinely available clinical data.

For our second research question whether, radiomics features can predict long-term disability progression in PwMS, the most generalisable model was the LGBM trained on harmonised data with the radiomics and clinical features subset. As seen in Figure 9, the most influential features constitute textural features from the WML and NAWM. The role of textural features in MS disease progression has been studied previously (Harrison et al., 2010; Herlidou-Même et al., 2003; Kassner & Thornhill, 2010; Loizou et al., 2010; Loizou, Kyriacou, et al., 2011; Loizou et al., 2015, 2020; Meier & Guttmann, 2003; J. Zhang et al., 2008) and our study further strengthens the notion that textural features can capture the diffuse pathological changes in these areas. The textural features extracted from WML tend to capture the heterogeneity and structural characteristics of the lesions, which can provide a non-invasive means of assessing lesion activity and overall burden, which is critical for MS disease progression (Y. Zhang et al., 2013). As studied before, the heterogeneity in voxel intensities corresponds to demyelination, axonal loss and inflammation in the WML (Barkovich, 2000; Y. Zhang et al., 2013). Furthermore, previous studies have shown that textural heterogeneity can act as relevant biomarkers to predict progression (Loizou, Murray, et al., 2011; Tozer et al., 2009). This could be due to the origin of the MRI signal from the endogenous protons which are affected by the structural changes at the microscopic level in pathology (NAWM and WML) causing magnetic resonance signal variation at the macroscopic scale (Y. Zhang et al., 2013).

The top predictor among the WML textural features in our study was GLCM Maximum Probability (WML), which shows an inverse relationship with disability progression. This feature essentially measures the most probable co-occurence of intensity values within an ROI (Haralick et al., 1973). In the case of WML, this would mean that a higher value of GLCM Maximum Probability would indicate a higher degree of homogeneity of intensity values, whereas lower values would indicate lower homogeneity or increased heterogeneity in the WML. Therefore, the more textural heterogeneity a lesion exhibits, the more demyelination and other microstructural changes occur within that lesion (Barkovich, 2000; Y. Zhang et al., 2013). The textural features extracted from the NAWM were also deemed as predictive features. They represent possible diffuse pathological changes such as gliosis or early demyelination, which are not visible to the naked eye on MRI. This is in line with the literature (Y. Zhang, 2012; Y. Zhang et al., 2009).

In addition to textural features, anatomical volumes such as left lateral ventricle volume and right amygdala volume were also deemed useful in our study. Although ventricular enlargement corresponds to brain atrophy, which corresponds further to disability progression, in our study, the ventricular enlargement exhibited a negative correlation with disability progression, unlike previous studies (Genovese et al., 2019; Jakimovski et al., 2020; Zivadinov et al., 2019). As shown in Appendix M, this negative correlation can be attributed to cases with advanced atrophy (high left lateral ventricle volume) being labelled as non-progressive, as their baseline disability score (EDSS_T0) is already high, leaving little room for measurable progression within the two-year follow-up period.

In contrast, a larger right amygdala volume was associated negatively with disability progression, pointing to the possible role of the limbic system in preserving cognitive and neurological function in MS. Lastly, the clinical features did not have as much of a higher influence as the others, but they remained important predictors nevertheless. The highest being the age at baseline exhibiting a positive correlation with disability progression (Kalnina et al., 2024).

For our last research question, i.e., whether short-term changes in MRI features can predict long-term disability progression, we used the longitudinal imaging prognostic approach in an attempt to capture temporal changes and exploit its use to make our models robust. We found that the BRFC trained on the non-harmonised radiomics feature subset achieved a PR AUC of 0.11 and ROC AUC of 0.69 on the test set. However, looking at the features and their corresponding SHAP values, we observed that the selected features, corresponding to the dynamic changes in lesion structure over time, exhibited a lower predictive power compared to the harmonised features selected in the baseline imaging approach. This could be due to the short temporal window between the baseline and follow up and a reduction in the dataset, which inhibits their capability to fully capture short term changes to explain long term disability progression in PwMS.

To further validate our findings and eliminate the risk of overfitting, the permutation results, presented in Appendix J, indicate that the permuted models’ performance was notably worse than the original models. This indicates that the predictive power of our models is driven by meaningful patterns in the data rather than random noise or spurious correlations. Furthermore, the poor performance of the permuted models validates the robustness of our approach, as any enhancement in prediction performance observed in the original models cannot be attributed to chance.

Interestingly, the combined prognostic approach did not yield a generalisable predictive performance, suggesting that the baseline imaging prognostic approach is sufficient to capture the majority of relevant information for predicting long-term progression. The integration of longitudinal imaging features may have introduced noise, diluting the predictive signal of the more robust baseline features. These results underscore the need for careful feature selection and refined temporal analysis to optimise combined approaches.

This study brings important advancements compared to the existing literature. By leveraging multicentric data from two centers with diverse MRI acquisition protocols, it enhances the generalisability of findings. The robust preprocessing pipeline, including super-resolution reconstruction and longitudinal ComBat harmonisation, attempted to ensure consistency in imaging data across sites and protocols. Additionally, the use of SHAP analysis provided interpretable insights into feature importance, offering a deeper understanding of the role of radiomics in predicting MS progression. While MRI, unlike computed tomography, is inherently non-quantitative, our study, similar to previous work (Lavrova et al., 2021), demonstrates the potential of radiomics features in capturing subtle pathological changes. The selection of radiomics features from the WML and NAWM, coupled with radiomics and clinical features, further enhances the promise radiomics holds to bridge the gap between radiological findings and clinical outcomes, also known as the clinico radiological paradox (Uitdehaag, 2018).

However, certain limitations must be acknowledged. The small number of worsening disability progression cases translated into a high class imbalance, which posed challenges despite the use of weighted adjustments. The reliance on reconstructed images without ground truth and the absence of T1-weighted sequences may have affected segmentation and feature quality. While initially, we performed ML analysis where DS2 was kept as a completely held out external set, the models tended to generalise poorly on it (see Appendix G). Even though longitudinal ComBat harmonisation attempts to mitigate scanner and site variability, its ability to preserve subtle predictive patterns and address batch effect warrants further validation. Finally, the retrospective design may introduce selection bias, limiting the generalisability of these findings to broader populations.

Future studies should address these limitations by incorporating larger, multicentric, balanced datasets with higher-resolution MRI and ground-truth labels. Expanding the temporal window for delta radiomics and integrating advanced imaging modalities, such as diffusion-weighted imaging, may enhance the predictive power of radiomics. Additionally, exploring the role of other clinical variables, such as disease modifying therapy, disease duration, alongside imaging biomarkers could provide a more comprehensive understanding of progression mechanisms in MS. Lastly, deep radiomics with pre-trained foundation models can be deployed to see whether a deep learning algorithm might be able to uncover patterns that the traditional ML algorithm with hand-crafted radiomics might have failed to capture.

## Conclusion

This study highlights the potential of FLAIR MRI-based radiomics combined with ML to predict long-term disability progression in PwMS. We demonstrated that models combining radiomics and clinical features outperforms clinical-only models. Furthermore, we found that radiomics features from WML and NAWM, and routine clinical features in baseline imaging prognostic approach emerged as predictors, reinforcing their diagnostic value. However, the longitudinal imaging approach demonstrated limited predictive power, emphasising the need for refined temporal analysis. Future work should address class imbalance, enhance feature quality, and explore advanced imaging modalities to further advance MS progression prediction.

## Supporting information

Appendix F Radiomics Quality Score (attached separately)

Appendix E Radiomics Check-list (CLEAR)

All Supplementary Tables and Figures

## Data Availability

The data supporting the findings of this study was obtained from the Rehabilitation and MS Center of Noorderhart in Pelt, Belgium and Zuyderland Medical Center in Sittard, the Netherlands. The study has been approved by the ethical commission of the University of Hasselt (CME2019/046) and the Medical Ethics Review Committee of Zuyderland and Zuyd University of Applied Sciences (METCZ20200167). The data is not publicly available due to privacy and confidentiality concerns.

## Acknowledgments

The authors thank Zohaib Salahuddin (The D-Lab, Department of Precision Medicine, GROW – Research Institute for Oncology and Reproduction, Maastricht University, Maastricht, Netherlands) for his valuable feedback and insights during the development of this study. We also acknowledge Raymond Hupperts (Academic MS Center Zuyd, Department of Neurology, Zuyderland Medical Center, Sittard-Geleen, Netherlands) for his guidance and support in shaping the clinical aspects of this work.

## Supplementary Materials

Added Seperately to the manuscript.

## Code Availability

The code used to conduct this study can be accessed on GitHub.

## Author Contributions

Conceptualisation, Hamza Khan, Liesbet M. Peeters, Henry C. Woodruff and Philippe Lambin; methodology, Hamza Khan, Diana L. Giraldo and Henry C.Woodruff; software, Hamza Khan and Diana L. Giraldo; validation, Hamza Khan, Diana L. Giraldo, Lorin Werthen-Brabants, Sina Amirrajab, and Edward De Brouwer; formal analysis, Hamza Khan and Diana L. Giraldo; investigation, Hamza Khan and Diana L. Giraldo; resources, Jan Sijbers, Oliver Gerlach, Veronica Popescu and Bart Van Wijmeersch; data curation, Hamza Khan and Diana L. Giraldo; writing—original draft preparation, Hamza Khan; writing—review and editing, Hamza Khan, Henry C Woodruff, Diana L. Giraldo, Lorin Werthen-Brabants, Shruti Atul Mali, Sina Amirrajab, Edward De Brouwer, Veronica Popescu, Bart Van Wijmeersch, Oliver Gerlach, Jan Sijbers, Liesbet M. Peeters and Philippe Lambin; visualisation, Hamza Khan; harmonisation, Hamza Khan, Shruti A Mali, and Diana L. Giraldo, supervision, Jan Sijbers, Henry C. Woodruff, Liesbet M. Peeters, and Philippe Lambin; project administration, Hamza Khan, Liesbet M. Peeters, and Philippe Lambin; funding acquisition, Liesbet M. Peeters, and Philippe Lambin. Raymond Hupperts (ZMC), Oliver Gerlach, Veronica Popescu and Bart Van Wijmeersch provided clinical insights. All authors have read and agreed to the published version of the manuscript.

## Funding

This research received funding from the Flemish Government under the “Onderzoeksprogramma Artificiële Intelligentie (AI) Vlaanderen” program, Stichting Multiple Sclerosis Research (19-1040 MS) and the Bijzonder OnderzoeksFonds (BOF19DOCMA10). Authors acknowledge financial support from the European Union’s Horizon research and innovation programme under grant agreement: ImmunoSABR n° 733008, CHAIMELEON n° 952172, EuCanImage n° 952103, IMI-OPTIMA n° 101034347, RADIOVAL (HORIZON-HLTH-2021-DISEASE-04-04) n°101057699, EUCAIM (DIGITAL-2022-CLOUD-AI-02) n°101100633, GLIOMATCH n° 101136670, AIDAVA (HORIZON-HLTH-2021-TOOL-06) n°101057062, REALM (HORIZON-HLTH-2022-TOOL-11) n° 101095435.

## Institutional Review Board Statement

The study has been approved by the ethical commission of the University of Hasselt (CME2019/046), and and the Medical Ethics Review Committee of Zuyderland and Zuyd University of Applied Sciences (METCZ20200167). No consent to participate was required, given the retrospective nature of the study. Furthermore, the images used were pseudonymised.

## Conflicts of interest

**Hamza Khan:** No conflict of interest or relevant disclosures to this study.

**Diana L. Giraldo:** No conflict of interest or relevant disclosures to this study.

**Henry C Woodruff:** Minority shares in the company Radiomics SA

**Lorin Werthen-Brabants:** No conflict of interest or relevant disclosures to this study.

**Shruti Atul Mali:** No conflict of interest or relevant disclosures to this study.

**Sina Amirrajab:** No conflict of interest or relevant disclosures to this study.

**Edward De Brouwer:** No conflict of interest or relevant disclosures to this study.

**Veronica Popescu:** No conflict of interest or relevant disclosures to this study.

**Bart Van Wijmeersch:** No conflict of interest or relevant disclosures to this study.

**Oliver Gerlach:** No conflict of interest or relevant disclosures to this study.

**Jan Sijbers:** No conflict of interest or relevant disclosures to this study.

**Liesbet M. Peeters:** No conflict of interest or relevant disclosures to this study.

**Philippe Lambin:** None related to the current manuscript; outside of current manuscript: grants/sponsored research agreements from Radiomics SA, Convert Pharmaceuticals SA and LivingMed Biotech srl. He received a presenter fee and/or reimbursement of travel costs/consultancy fee (in cash or in kind) from Astra Zeneca, BHV srl & Roche. PL has/had minority shares in the companies Radiomics SA, Convert pharmaceuticals SA, Comunicare SA, LivingMed Biotech srl and Bactam srl. PL is co-inventor of two issued patents with royalties on radiomics (PCT/NL2014/050248 and PCT/NL2014/050728), licensed to Radiomics SA; one issued patent on mtDNA (PCT/EP2014/059089), licensed to ptTheragnostic/DNAmito; one granted patent on LSRT (PCT/ P126537PC00, US patent No. 12,102,842), licensed to Varian; one issued patent on Radiomic signature of hypoxia (U.S. Patent 11,972,867), licensed to a commercial entity; one issued patent on Prodrugs (WO2019EP64112) without royalties; one non-issued, non-licensed patents on Deep Learning-Radiomics (N2024889) and three non-patented inventions (softwares) licensed to ptTheragnostic/DNAmito, Radiomics SA and Health Innovation Ventures). Philippe Lambin confirms that none of the above entities were involved in the preparation of this paper.

## Notes

### Funding Statement

This research received funding from the Flemish Government under the Onderzoeksprogramma Artificiele Intelligentie (AI) Vlaanderen program, Stichting Multiple Sclerosis Research (19-1040 MS), and the Bijzonder OnderzoeksFonds (BOF19DOCMA10). Authors acknowledge financial support from the European Union's Horizon research and innovation programme under grant agreements: ImmunoSABR n[deg] 733008, CHAIMELEON n[deg] 952172, EuCanImage n[deg] 952103, IMI-OPTIMA n[deg] 101034347, RADIOVAL (HORIZON-HLTH-2021-DISEASE-04-04) n[deg] 101057699, EUCAIM (DIGITAL-2022-CLOUD-AI-02) n[deg] 101100633, GLIOMATCH n[deg] 101136670, AIDAVA (HORIZON-HLTH-2021-TOOL-06) n[deg] 101057062, REALM (HORIZON-HLTH-2022-TOOL-11) n[deg] 101095435.

### Author Declarations

The Ethics Committee of the University of Hasselt (CME2019/046) and the Medical Ethics Review Committee of Zuyderland and Zuyd University of Applied Sciences (METCZ20200167) provided ethical approval for this study. No consent to participate was required due to the retrospective nature of the research. All images were pseudonymized to ensure participant confidentiality.

