## Appendix F Radiomics Quality Score (attached separately) for "Leveraging Hand-Crafted Radiomics on Multicenter FLAIR MRI for Predicting Disability Progression in People with Multiple Sclerosis"

Image protocol quality - well-documented image protocols (for example, contrast, slice thickness, energy, etc.) and/or usage of public image protocols allow reproducibility/replicability

- [x] protocols well documented
- [ ] public protocol used
- [ ] none

Multiple segmentations - possible actions are: segmentation by different physicians/algorithms/software, perturbing segmentations by (random) noise, segmentation at different breathing cycles. Analyse feature robustness to segmentation variabilities

- ( ) yes
- (•) no

Phantom study on all scanners - detect inter-scanner differences and vendor-dependent features. Analyse feature robustness to these sources of variability

- ( ) yes
- (•) no

Imaging at multiple time points - collect images of individuals at additional time points. Analyse feature robustness to temporal variabilities (for example, organ movement, organ expansion/shrinkage)

- (•) yes
- ( ) no

Feature reduction or adjustment for multiple testing - decreases the risk of overfitting. Overfitting is inevitable if the number of features exceeds the number of samples. Consider feature robustness when selecting features

- (•) Either measure is implemented
- ( ) Neither measure is implemented

Multivariable analysis with non radiomics features (for example, EGFR mutation) - is expected to provide a more holistic model. Permits correlating/inferencing between radiomics and non radiomics features

- (•) yes
- ( ) no

Detect and discuss biological correlates - demonstration of phenotypic differences (possibly associated with underlying gene–protein expression patterns) deepens understanding of radiomics and biology

- ( ) yes
- (•) no

Cut-off analyses - determine risk groups by either the median, a previously published cut-off or report a continuous risk variable. Reduces the risk of reporting overly optimistic results

- ( ) yes

- [x] no

Discrimination statistics - report discrimination statistics (for example, C-statistic, ROC curve, AUC) and their statistical significance (for example, p-values, confidence intervals). One can also apply resampling method (for example, bootstrapping, cross-validation)

- [x] a discrimination statistic and its statistical significance are reported
- [x] a resampling method technique is also applied
- [ ] none

Calibration statistics - report calibration statistics (for example, Calibration-in-the-large/slope, calibration plots) and their statistical significance (for example, P-values, confidence intervals). One can also apply resampling method (for example, bootstrapping, cross-validation)

- [ ] a calibration statistic and its statistical significance are reported
- [ ] a resampling method technique is applied
- [x] none

Prospective study registered in a trial database - provides the highest level of evidence supporting the clinical validity and usefulness of the radiomics biomarker

- [ ] yes
- [x] no

Validation - the validation is performed without retraining and without adaptation of the cut-off value, provides crucial information with regard to credible clinical performance

- [ ] No validation
- [ ] validation is based on a dataset from the same institute
- [ ] validation is based on a dataset from another institute
- [x] validation is based on two datasets from two distinct institutes
- [ ] the study validates a previously published signature
- [ ] validation is based on three or more datasets from distinct institutes

Comparison to 'gold standard' - assess the extent to which the model agrees with/is superior to the current 'gold standard' method (for example, TNM-staging for survival prediction). This comparison shows the added value of radiomics

- [ ] yes
- [x] no

Potential clinical utility - report on the current and potential application of the model in a clinical setting (for example, decision curve analysis).

- [x] yes
- [ ] no

Cost-effectiveness analysis - report on the cost-effectiveness of the clinical application (for example, QALYs generated)

- [ ] yes
- [x] no

Open science and data - make code and data publicly available. Open science facilitates knowledge transfer and reproducibility of the study

- [ ] scans are open source
- [ ] region of interest segmentations are open source
- [x] the code is open sourced
- [ ] radiomics features are calculated on a set of representative ROIs and the calculated features and representative ROIs are open source

Total score **15** (41.67%)
