## Appendix E Radiomics Check-list (CLEAR) for "Leveraging Hand-Crafted Radiomics on Multicenter FLAIR MRI for Predicting Disability Progression in People with Multiple Sclerosis"

**Electronic Supplementary Material S2:** CLEAR checklist without explanations

| Section | No. | Item | Yes | No | n/a | Page |
| --- | --- | --- | --- | --- | --- | --- |
| Title |  |  |  |  |  |  |
|  | 1 | Relevant title, specifying the radiomic methodology | X | ☐ | ☐ | - |
| Abstract |  |  |  |  |  |  |
|  | 2 | Structured summary with relevant information | X | ☐ | ☐ | - |
| Keywords |  |  |  |  |  |  |
|  | 3 | Relevant keywords for radiomics | X | ☐ | ☐ | - |
| Introduction |  |  |  |  |  |  |
|  | 4 | Scientific or clinical background | X | ☐ | ☐ | 1 |
|  | 5 | Rationale for using a radiomic approach | X | ☐ | ☐ | 2 |
|  | 6 | Study objective(s) | X | ☐ | ☐ | 2 |
| Method |  |  |  |  |  |  |
| *Study Design* | 7 | Adherence to guidelines or checklists (e.g., CLEAR checklist) | X | ☐ | ☐ | 11 |
|  | 8 | Ethical details (e.g., approval, consent, data protection) | X | ☐ | ☐ | 5 |
|  | 9 | Sample size calculation | ☐ | ☐ | X |  |
|  | 10 | Study nature (e.g., retrospective, prospective) | X | ☐ | ☐ | 6 |
|  | 11 | Eligibility criteria | X | ☐ | ☐ | 5 |
|  | 12 | Flowchart for technical pipeline | X | ☐ | ☐ | 7 |
| *Data* | 13 | Data source (e.g., private, public) | X | ☐ | ☐ | 5 |
|  | 14 | Data overlap | X | ☐ | ☐ | 5 |
|  | 15 | Data split methodology | X | ☐ | ☐ | 9 |
|  | 16 | Imaging protocol (i.e., image acquisition and processing) | X | ☐ | ☐ | 5 |
|  | 17 | Definition of non-radiomic predictor variables | X | ☐ | ☐ | 5 |
|  | 18 | Definition of the reference standard (i.e., outcome variable) | X | ☐ | ☐ | 6 |
| *Segmentation* | 19 | Segmentation strategy | X | ☐ | ☐ | 7 |
|  | 20 | Details of operators performing segmentation | ☐ | X | ☐ |  |
| *Pre-processing* | 21 | Image pre-processing details | X | ☐ | ☐ | 5 |
|  | 22 | Resampling method and its parameters (Super resolution reconstruction using PRETTIER) | ☐ | X | ☐ | 6 |
|  | 23 | Discretization method and its parameters | X | ☐ | ☐ | 8 |
|  | 24 | Image types (e.g., original, filtered, transformed) | ☐ | ☐ | X |  |
| *Feature extraction* | 25 | Feature extraction method | X | ☐ | ☐ | 8 |
|  | 26 | Feature classes | X | ☐ | ☐ | 8 |
|  | 27 | Number of features | X | ☐ | ☐ | 8 |
|  | 28 | Default configuration statement for remaining parameters | ☐ | ☐ | ☐ |  |
| *Data preparation* | 29 | Handling of missing data | ☐ | ☐ | X |  |
|  | 30 | Details of class imbalance | ☐ | ☐ | X |  |
|  | 31 | Details of segmentation reliability analysis | ☐ | X | ☐ |  |
|  | 32 | Feature scaling details (e.g., normalization, standardization) | X | ☐ | ☐ | 8 |
|  | 33 | Dimension reduction details | X | ☐ | ☐ | 10 |
| *Modeling* | 34 | Algorithm details | X | ☐ | ☐ | 11 |
|  | 35 | Training and tuning details | X | ☐ | ☐ | 11 |
|  | 36 | Handling of confounders | ☐ | ☐ | X |  |
|  | 37 | Model selection strategy | X | ☐ | ☐ | 11 |
| *Evaluation* | 38 | Testing technique (e.g., internal, external) (Note – data from two centers was mixed) | X | ☐ | ☐ | 9 |
|  | 39 | Performance metrics and rationale for choosing | X | ☐ | ☐ | 11 |
|  | 40 | Uncertainty evaluation and measures (e.g., confidence intervals) | ☐ | X | ☐ |  |
|  | 41 | Statistical performance comparison (e.g., DeLong’s test) | X | ☐ | ☐ | 11 |
|  | 42 | Comparison with non-radiomic and combined methods | X | ☐ | ☐ | 10 |
|  | 43 | Interpretability and explainability methods | X | ☐ | ☐ | 11 |
| Results |  |  |  |  |  |  |
|  | 44 | Baseline demographic and clinical characteristics | X | ☐ | ☐ | 13 |
|  | 45 | Flowchart for eligibility criteria | ☐ | ☐ | X |  |
|  | 46 | Feature statistics (e.g., reproducibility, feature selection) | X | ☐ | ☐ | 15 |
|  | 47 | Model performance evaluation | X | ☐ | ☐ | 16 |
|  | 48 | Comparison with non-radiomic and combined approaches | X | ☐ | ☐ | 18 |
| Discussion |  |  |  |  |  |  |
|  | 49 | Overview of important findings | X | ☐ | ☐ | 21 |
|  | 50 | Previous works with differences from the current study | X | ☐ | ☐ | 22 |
|  | 51 | Practical implications | X | ☐ | ☐ | 23 |
|  | 52 | Strengths and limitations (e.g., bias and generalizability issues) | X | ☐ | ☐ | 23 |
| Open Science |  |  |  |  |  |  |
| *Data availability* | 53 | Sharing images along with segmentation data [n/e] | ☐ | X | ☐ |  |
|  | 54 | Sharing radiomic feature data | ☐ | X | ☐ |  |
| *Code availability* | 55 | Sharing pre-processing scripts or settings | X | ☐ | ☐ |  |
|  | 56 | Sharing source code for modeling | X | ☐ | ☐ |  |
| *Model availability* | 57 | Sharing final model files | ☐ | X | ☐ |  |
|  | 58 | Sharing a ready-to-use system [n/e] | ☐ | X | ☐ |  |

**Yes**, details provided; **No**, details not provided; **n/e**, not essential; **n/a**, not applicable

Note: Use the checklist in conjunction with the main text for clarification of all items. Fill the “Page” column with the related page number where the information is provided.
