## Supplementary material for "Leveraging Hand-Crafted Radiomics on Multicenter FLAIR MRI for Predicting Disability Progression in People with Multiple Sclerosis": All Supplementary Tables and Figures

### Supplementary Information

Appendix A: T2-W FLAIR MRI acquisition details

For DS1-Pelt:

| N sessions | Scanner | Protocol | Acquisition type | Images per session | Repetition time | Echo time | Inversion time | Slice thickness | Slice spacing | Pixel spacing |
|---|---|---|---|---|---|---|---|---|---|---|
| 487 | Philips Achieva 1.5T | A | 2D | 2 | 6000 ms | 120 ms | 2000 ms | 5 mm | 6 mm | 0.72 - 0.90 mm |
| 64 | Philips Achieva 1.5T | B | 3D | 3 | 4800 ms | 321 - 363 ms | 1660 ms | 3 mm | 3 mm | 0.98 mm |
| 79 | Philips Achieva 1.5T | C/CsT1 | 3D | 1 | 4800 ms | 323 - 356 ms | 1660 ms | 1.2 mm | 0.6 mm | 0.98 mm |

For DS2-Zuyderland:

| N sessions | Scanner | Protocol | Acquisition type | Images per session | Repetition time | Echo time | Inversion time | Slice thickness | Slice spacing | Pixel spacing |
|---|---|---|---|---|---|---|---|---|---|---|
| 9 | Philips Achieva 1.5T | D | 2D | 1 | 11000 ms | 140 ms | 2800 ms | 5 mm | 6 mm | 0.9 mm |
| 73 | Philips Achieva 1.5T | A | 2D | 2 | 9000 - 11000 ms | 140 ms | 2800 ms | 5 mm | 6 mm | 0.8 - 0.9 mm |
| 2 | Philips Achieva 1.5T | C | 3D | 1 | 4800 ms | 319 ms | 1660 ms | 1 mm | 1 mm | 0.96 mm |

| | | | | | | | | | | |
|---|---|---|---|---|---|---|---|---|---|---|
| 2 | Philips Achieva 3T | D | 2D | 1 | 11000 ms | 125 ms | 2800 ms | 5 mm | 5.5 mm | 0.45 mm |
| 1 | Philips Ingenia 3T | C | 3D | 1 | 4800 ms | 300 ms | 1650 ms | 0.9 mm | 0.9 mm | 0.24 mm |
| 2 | SIEMENS Aera 1.5T | A | 2D | 2 | 9000 ms | 86 - 90 ms | 2500 ms | 3 - 5mm | 3.9 - 6.5 mm | 0.72 - 0.73 mm |
| 15 | SIEMENS Avanto 1.5T | D | 2D | 1 | 8000 - 11900 ms | 102 - 125 ms | 2500 ms | 3 - 5 mm | 3 - 7 mm | 0.45 - 1 mm |
| 21 | SIEMENS Avanto 1.5T | A | 2D | 2 | 6000 - 11900 ms | 102 - 125 ms | 2500 ms | 3 - 5 mm | 3 - 7 mm | 0.45 - 1 mm |
| 1 | SIEMENS Avanto_fit 1.5T | D | 2D | 1 | 9000 ms | 85 ms | 2500 ms | 4 mm | 4.4 mm | 0.72 mm |
| 35 | SIEMENS Avanto_fit 1.5T | A | 2D | 2 | 6000 - 9000 ms | 85 - 107 ms | 2020 - 2500 ms | 4 - 5 mm | 4.4 - 5.5 mm | 0.45 - 0.9 mm |
| 1 | SIEMENS Avanto_fit 1.5T | CsT1 | 3D | 1 | 5000 ms | 381 ms | 1800 ms | 1 mm | 1 mm | 1 mm |
| 1 | SIEMENS MAGNETOM EXPERT plus 0.95T | A | 2D | 2 | 9000 ms | 105 ms | 2223 ms | 5 mm | 6.5 mm | 0.98 mm |
| 2 | SIEMENS Skyra 3T | D | 2D | 1 | 9400 ms | 78 - 112 ms | 2500 ms | 3 mm | 3 mm | 1 mm |
| 10 | SIEMENS Skyra 3T | A | 2D | 2 | 9000 ms | 81 ms | 2500 ms | 4 mm | 4.4 - 5.2 mm | 0.72 mm |
| 7 | SIEMENS Skyra 3T | CsT1 | 3D | 1 | 5000 ms | 384 - 388 ms | 1800 ms | 0.9 - 1 mm | 0.9 - 1 mm | 0.9 - 1 mm |
| 2 | SIEMENS Symphony | A | 2D | 2 | 9000 ms | 106 - 128 ms | 2500 ms | 3 - 5 mm | 3.9 - 6.5 mm | 0.45 - 0.9 mm |

| | |
|---|---|
| | 1.5T |

Appendix B: Anatomical brain structures segmented by SAMSEG

| Number | Feature |
|---|---|
| 1 | left cerebral cortex volume |
| 2 | right cerebral cortex volume |
| 3 | left cerebral white matter volume |
| 4 | right cerebral white matter volume |
| 5 | left cerebellum cortex volume |
| 6 | right cerebellum cortex volume |
| 7 | left cerebellum white matter volume |
| 8 | right cerebellum white matter volume |
| 9 | right amygdala volume |
| 10 | left amygdala volume |
| 11 | right hippocampus volume |
| 12 | left hippocampus volume |
| 13 | left accumbens area volume |
| 14 | right accumbens area |
| 15 | left putamen volume |
| 16 | right putamen volume |
| 17 | right pallidum volume |
| 18 | left left pallidum volume |
| 19 | left caudate volume |
| 20 | right caudate volume |
| 21 | right thalamus volume |

| | |
|---|---|
| 22 | left thalamus volume |
| 23 | right choroid plexus volume |
| 24 | left choroid plexus volume |
| 25 | right ventral diencephalon volume |
| 26 | left ventral diencephalon volume |
| 27 | right inferior lateral ventricle volume |
| 28 | left inferior lateral ventricle volume |
| 29 | fourth ventricle volume |
| 30 | third ventricle volume |
| 31 | left lateral ventricle volume |
| 32 | right lateral ventricle volume |
| 33 | cerebrospinal fluid volume |
| 34 | brain stem volume |
| 35 | cerebral grey matter cortex volume |
| 36 | cerebral white matter volume |
| 37 | ventricle volume |
| 38 | total white matter lesion volume |
| 39 | intra cranial volume |
| 40 | fifth ventricle volume |
| 41 | unknowns volume |

Appendix C: Features selected for all prognostic approaches and feature subsets

| **Approach** | **Harmonisation** | **Feature Subset** | **Total Features** | **Non inter-correlated Features** | **N** | **Selected Features** |
|---|---|---|---|---|---|---|
| Clinical | Not applicable | Clinical only | 3 | - | 3 | EDSS_T0, clinical age in years, gender (female) |
| Baseline imaging | Harmonised | Radiomics volume features | 41 | 41 | 4 | right hippocampus volume, left lateral ventricle volume, right thalamus volume, right amygdala volume |
| | | Radiomics features without volumes | 200 | 103 | 6 | GLRLM run variance (WML), firstorder kurtosis (NAWM), GLCM maximum probability (WML), GLSZM large area low gray level emphasis (WML), shape minor axis length (NAWM), GLDM dependence non uniformity (NAWM) |
| | | Radiomics | 10 (selected features from radiomics volume features and radiomics features without volumes) | 10 (selected features from radiomics volume features and radiomics features without volumes) | 7 | GLRLM run variance (WML), GLCM maximum probability (WML), firstorder kurtosis (NAWM), left lateral ventricle volume, GLDM dependence non uniformity (NAWM), right amygdala volume, GLSZM large area low gray level emphasis (WML) |
| | | Radiomics and clinical features | 13 (selected features from radiomics | 13 (selected features from radiomics along with clinical features) | 10 | GLRLM run variance (WML), GLCM maximum probability (WML), firstorder kurtosis (NAWM), left lateral ventricle volume, GLDM dependence |

| | | | | | | |
|---|---|---|---|---|---|---|
| | | | along with clinical features) | | | non uniformity (NAWM), right amygdala volume, GLSZM large area low gray level emphasis (WML), EDSS_T0, clinical age in years, gender (female) |
| | Non harmonised | Radiomics volume features | 41 | 41 | 12 | left lateral ventricle volume, right accumbens area volume, left thalamus volume, right thalamus volume, left left pallidum volume, right amygdala volume, left choroid plexus volume, left caudate volume, left ventral diancephalon volume, left accumbens area volume, left hippocampus volume, right inferior lateral ventricle volume |
| | | Radiomics features without volumes | 200 | 98 | 19 | GLRLM run variance (WML), firstorder minimum (WML), first order 10 percentile (NAWM), GLCM cluster shade (WML), shape minor axis length (NAWM), GLDM large dependence high gray level emphasis (WML), GLDM dependence entropy (NAWM), shape maximum 2D diameter slice (WML), shape maximum 2D diameter column (NAWM), shape major axis length (WML), shape sphericity (NAWM), GLRLM run entropy (NAWM), GLCM joint energy (WML), shape surface volume ratio (NAWM), GLCM maximum probability (WML), shape flatness (NAWM), firstorder mean (WML), |

| | | | | | | |
|---|---|---|---|---|---|---|
| | | | | | | GLSZM size zone non uniformity (WML), GLCM joint entropy (WML) |
| | | Radiomics | 31 (selected features from radiomics volume features and radiomics features without volumes) | 31 (selected features from radiomics volume features and radiomics features without volumes) | 10 | GLRLM run variance (WML), left thalamus volume, left lateral ventricle volume, firstorder minimum (WML), right accumbens area volume, right thalamus volume, left left pallidum volume, right amygdala volume, GLSZM size zone non uniformity (WML), shape minor axis length (NAWM) |
| | | Radiomics and clinical features | 34 (selected features from radiomics along with clinical features) | 34 (selected features from radiomics along with clinical features) | 13 | GLRLM run variance (WML), left thalamus volume, left lateral ventricle volume, firstorder minimum (WML), right amygdala volume, right accumbens area volume, right thalamus volume, left left pallidum volume, GLSZM size zone non uniformity (WML), shape minor axis length (NAWM), EDSS_T0, clinical age in years, gender (female) |
| Longitudinal imaging | Harmonised | Radiomics volume features | 41 | 38 | 1 | delta right cerebellum cortex volume |
| | | Radiomics features without volumes | 200 | 179 | 1 | delta GLCM difference entropy (WML) |
| | | Radiomics | 2 (selected features | 2 (selected features from | 2 | delta Right cerebellum cortex volume, delta GLCM difference |

| | | | | | | |
|---|---|---|---|---|---|---|
| | | | from radiomics volume features and radiomics features without volumes) | radiomics volume features and radiomics features without volumes) | | entropy (WML) |
| | | Radiomics and clinical features | 5 (selected features from radiomics along with clinical features) | 5 (selected features from radiomics along with clinical features) | 5 | delta right cerebellum cortex volume, delta GLCM difference entropy (WML), EDSS_T0, delta clinical age in years, gender (female) |
| | Non harmonised | Radiomics volume features | 41 | 38 | 1 | delta left choroid plexus volume |
| | | Radiomics features without volumes | 200 | 178 | 2 | delta GLCM difference entropy (WML), delta GLDM gray level non uniformity (WML) |
| | | Radiomics | 3 (selected features from radiomics volume features and radiomics features without volumes) | 3 (selected features from radiomics volume features and radiomics features without volumes) | 3 | delta GLCM difference entropy (WML), delta GLDM gray level non uniformity (WML), delta left choroid plexus volume |
| | | Radiomics and clinical | 6 (selected features | 6 (selected features from | 6 | delta GLCM difference entropy (WML), delta GLDM gray level |

| | | | | | | |
|---|---|---|---|---|---|---|
| | | features | from radiomics along with clinical features) | radiomics along with clinical features) | | non uniformity (WML), delta left choroid plexus volume, EDSS_T0, delta clinical age in years, gender (female) |
| Combined | Harmonised | Radiomics volume features | 82 | 77 | 5 | left thalamus volume, delta Left cerebellum cortex volume, delta right hippocampus volume, delta right thalamus volume, right thalamus volume |
| | | Radiomics features without volumes | 400 | 222 | 1 | delta GLSZM_large area high gray level emphasis (NAWM) |
| | | Radiomics | 6 (selected features from radiomics volume features and radiomics features without volumes) | 6 (selected features from radiomics volume features and radiomics features without volumes) | 6 | left thalamus volume, delta Left cerebellum cortex volume, delta right hippocampus volume, delta right thalamus volume, right thalamus volume, delta GLSZM_large area high gray level emphasis (NAWM) |
| | | Radiomics and clinical features | 8 (selected features from radiomics along with clinical features) | 8 (selected features from radiomics along with clinical features) | 8 | left thalamus volume, delta Left cerebellum cortex volume, delta right hippocampus volume, delta right thalamus volume, right thalamus volume, delta GLSZM_large area high gray level emphasis (NAWM), clinical_age, delta clinical_age, gender (female) |
| | Non harmonised | Radiomics volume | 82 | 77 | 6 | delta right caudate volume, right thalamus volume, delta |

| | | | | | | |
|---|---|---|---|---|---|---|
| | | features | | | | left cerebellum cortex volume, left thalamus volume, delta right thalamus volume, brain stem volume |
| | | Radiomics features without volumes | 400 | 216 | 2 | delta GLDM large dependence high gray level emphasis (NAWM),<br>delta GLSZM gray level non uniformity (NAWM) |
| | | Radiomics | 8 (selected features from radiomics volume features and radiomics features without volumes) | 8 (selected features from radiomics volume features and radiomics features without volumes) | 7 | delta GLSZM gray level non uniformity (NAWM), left thalamus volume, delta GLDM large dependence high gray level emphasis (NAWM), right thalamus volume, brain stem volume, delta right thalamus volume, delta right caudate volume |
| | | Radiomics and clinical features | 11 (selected features from radiomics along with clinical features) | 11 (selected features from radiomics along with clinical features) | 11 | delta GLSZM gray level non uniformity (NAWM), left thalamus volume, delta GLDM large dependence high gray level emphasis (NAWM), right thalamus volume, brain stem volume, delta right thalamus volume, delta right caudate volume, EDSS_T0, clinical age in years, delta clinical age in years, gender (female) |

Appendix D : Performances of best models trained per feature subset

| **Approach** | **Harmonisation** | **Feature Subset** | **Best Model** | **Validation PR** | **Validation AUC** | **Test PR** | **Test AUC** |
|---|---|---|---|---|---|---|---|
| Clinical | Not applicable | Clinical only | LGBM | 0.16 | 0.65 | 0.08 | 0.6 |
| Baseline imaging | Harmonised (LongCombat) | Radiomics volume features | LOGIT | 0.13 | 0.68 | 0.08 | 0.56 |
| | | Radiomics without volumes | LOGIT | 0.2 | 0.64 | 0.12 | 0.74 |
| | | Radiomics | BRFC | 0.14 | 0.76 | 0.13 | 0.67 |
| | | Radiomics and clinical | LGBM | 0.25 | 0.65 | 0.2 | 0.64 |
| | Non harmonised | Radiomics volume features | LGBM | 0.13 | 0.59 | 0.09 | 0.62 |
| | | Radiomics without volumes | BRFC | 0.11 | 0.61 | 0.16 | 0.74 |
| | | Radiomics | BRFC | 0.18 | 0.72 | 0.2 | 0.63 |
| | | Radiomics and clinical | BRFC | 0.22 | 0.69 | 0.13 | 0.74 |
| Longitudinal imaging | Harmonised (LongCombat) | Radiomics volume features | BRFC | 0.41 | 0.66 | 0.25 | 0.48 |

| | | | | | | | |
|---|---|---|---|---|---|---|---|
| | | Radiomics without volumes | LOGIT | 0.30 | 0.52 | 0.06 | 0.49 |
| | | Radiomics | LOGIT | 0.30 | 0.51 | 0.05 | 0.38 |
| | | Radiomics and clinical | LOGIT | 0.30 | 0.51 | 0.06 | 0.41 |
| | Non harmonised | Radiomics volume features | BRFC | 0.08 | 0.73 | 0.27 | 0.67 |
| | | Radiomics without volumes | BRFC | 0.32 | 0.8 | 0.14 | 0.66 |
| | | Radiomics | BRFC | 0.32 | 0.78 | 0.11 | 0.69 |
| | | Radiomics and clinical | LOGIT | 0.31 | 0.64 | 0.08 | 0.53 |
| Combined | Harmonised (LongCombat) | Radiomics volume features | LGBM | 0.36 | 0.81 | 0.06 | 0.45 |
| | | Radiomics without volumes | LGBM | 0.21 | 0.44 | 0.07 | 0.52 |
| | | Radiomics | LOGIT | 0.54 | 0.9 | 0.06 | 0.41 |
| | | Radiomics and clinical | BRFC | 0.46 | 0.68 | 0.06 | 0.45 |
| | Non harmonised | Radiomics volume features | LGBM | 0.33 | 0.72 | 0.05 | 0.38 |

| | | | | | | | |
|---|---|---|---|---|---|---|---|
| | | Radiomics without volumes | BRFC | 0.20 | 0.42 | 0.05 | 0.41 |
| | | Radiomics | LGBM | 0.31 | 0.65 | 0.05 | 0.30 |
| | | Radiomics and clinical | LGBM | 0.53 | 0.91 | 0.06 | 0.44 |

Appendix E Radiomics Check-list (CLEAR) - attached seperately

Check_list_Radiomics_only.docx

Appendix F Radiomics Quality Score (attached separately)

RQS - Radiomics.world.pdf

Appendix G Model Results with keeping ZMC as external

| Approach | Harmonisation | Feature Subset | Best Model | Validation PR AUC | Validation ROC AUC | Test PR AUC | Test ROC AUC |
|---|---|---|---|---|---|---|---|
| Clinical | Not applicable | Clinical only | LOGIT | 0.1 | 0.65 | 0.09 | 0.32 |
| Baseline imaging | Harmonised (LongCombat) | Radiomics | LOGIT | 0.1 | 0.74 | 0.13 | 0.47 |
| | Non harmonised | Radiomics | LGBM | 0.16 | 0.6 | 0.13 | 0.44 |
| Longitudinal imaging | Harmonised (LongCombat) | Radiomics | LOGIT | 0.15 | 0.73 | 0.11 | 0.48 |
| | Non harmonised | Radiomics features without volume | BRFC | 0.17 | 0.63 | 0.08 | 0.45 |
| Combined | Harmonised (LongCombat) | Radiomics and clinical | LOGIT | 0.18 | 0.59 | 0.09 | 0.46 |
| | Non harmonised | Radiomics and clinical | BRFC | 0.17 | 0.53 | 0.08 | 0.45 |

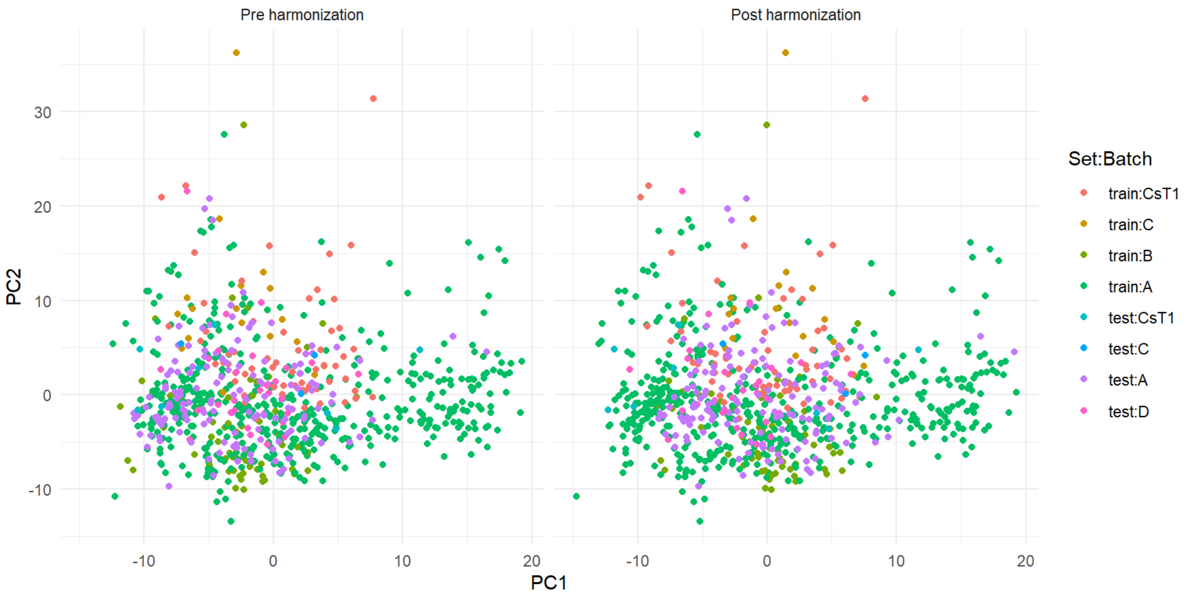

***Appendix H: Principal Component Analysis (PCA) Before and After Longitudinal ComBat Harmonisation.***
*The PCA plots illustrate the distribution of radiomic features across different batches and datasets (train and test) before (left) and after (right) harmonisation using longitudinal ComBat.*

**Appendix I: Best Models Parameters**

| Approach | Harmonisation | Feature Subset | Best Model | Model Parameters |
|---|---|---|---|---|
| Clinical | Not applicable | Clinical only | LGBM | weight_class_1: 2.861272625103398<br>n_estimators: 149<br>max_depth: 7<br>learning_rate: 0.04500730948996164<br>num_leaves: 26 |
| Baseline imaging | Harmonised (Long Combat) | Radiomics and clinical | LGBM | weight_class_1: 2.724695499698599<br>n_estimators: 158<br>max_depth: 10<br>learning_rate: 0.055581462804548766<br>num_leaves: 20 |
| Longitudinal imaging | Non Harmonised | Radiomics | BRFC | weight_class_1: 2.3572082945950172<br>n_estimators: 36<br>max_depth: 4<br>min_samples_split: 4<br>min_samples_leaf: 2 |

Appendix J: Permutation results achieved on the best approach and feature subset combination

| **Approach** | **Harmonisation** | **Feature Subset** | **Best Model** | **Validation PR AUC** | **Validation ROC AUC** |
|---|---|---|---|---|---|
| Clinical | Not applicable | Clinical only | LGBM | 0.08 ± 0.03 | 0.45 ± 0.06 |
| Baseline imaging | Harmonised (Long Combat) | Radiomics and clinical | LGBM | 0.007 ± 0.01 | 0.48 ± 0.05 |
| Longitudinal imaging | Non Harmonised | Radiomics | BRFC | 0.03 ± 0.04 | 0.46 ± 0.1 |

Appnedix K: Number of Radiomics Features Per Class

| **Feature Class** | **Number of Features Extracted per ROI** |
|---|---|
| shape | 14 |
| first-order statistics (FO) | 18 |
| gray-level co-occurrence matrix (GLCM) | 22 |
| gray-level run length matrix (GLRLM) | 16 |
| gray-level size zone matrix (GLSZM) | 16 |
| gray-level dependence matrix (GLDM) | 14 |

#### Appendix L: Distribution plots of disability progression in selected features of both Baseline and Longitudinal prognostic approach

Baseline imaging features plots.

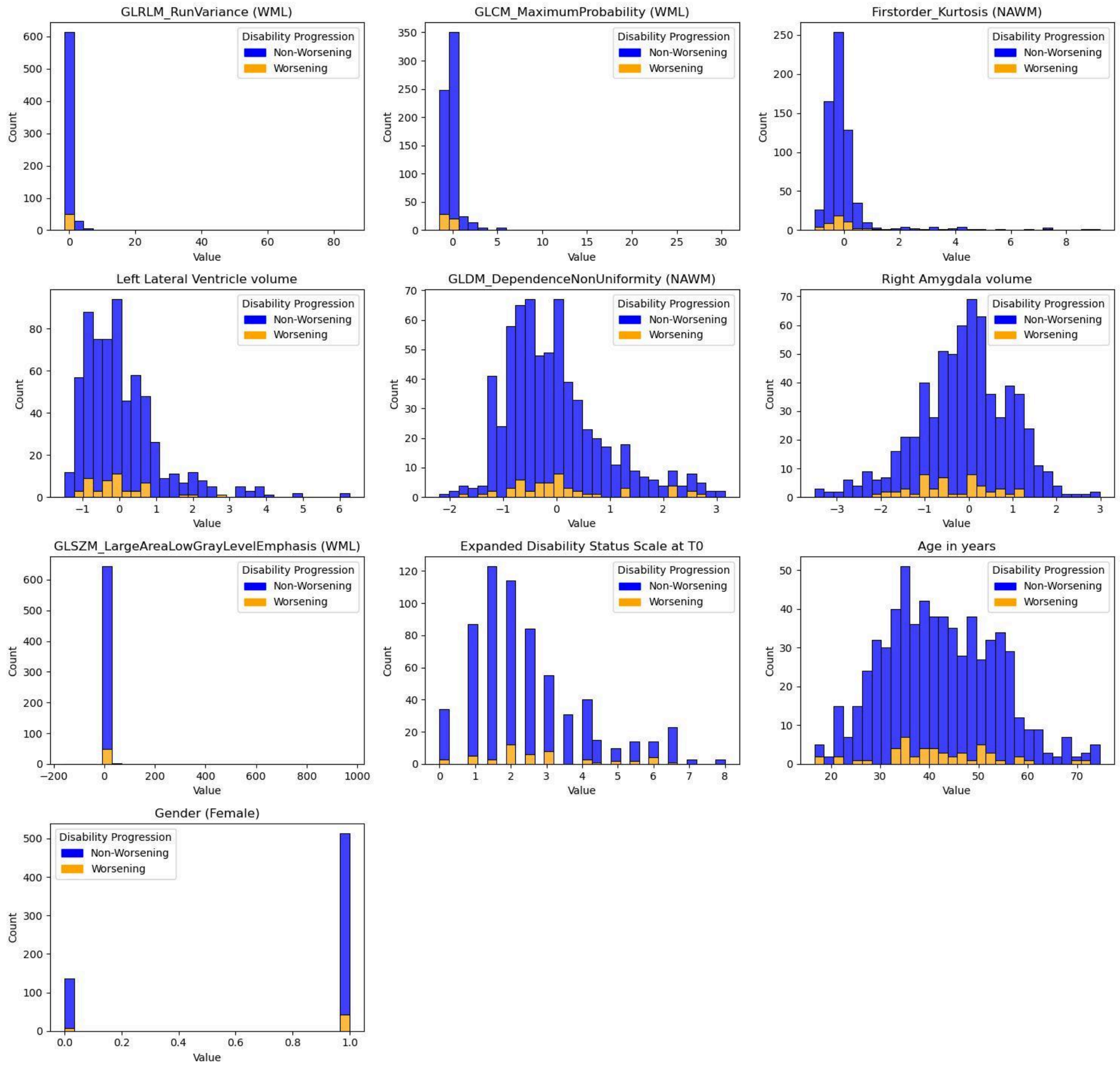

Longitudinal imaging Features Plots:

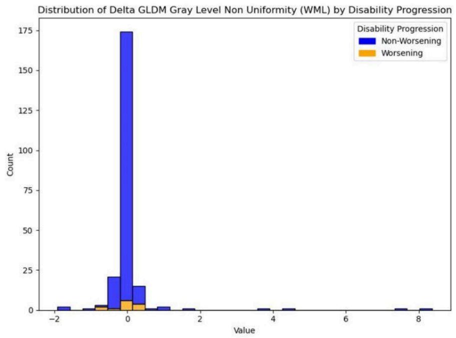
Distribution of Delta GLDM Gray Level Non Uniformity (WML) by Disability Progression
Disability Progression
Non-Worsening
Worsening
Count
Value

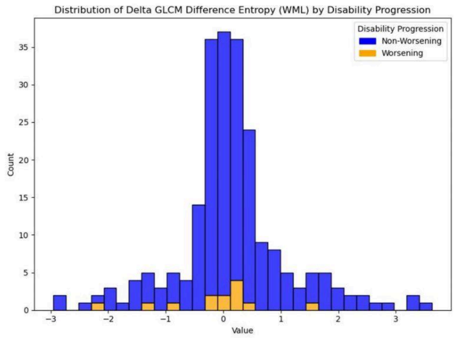
Distribution of Delta GLCM Difference Entropy (WML) by Disability Progression
Disability Progression
Non-Worsening
Worsening
Count
Value

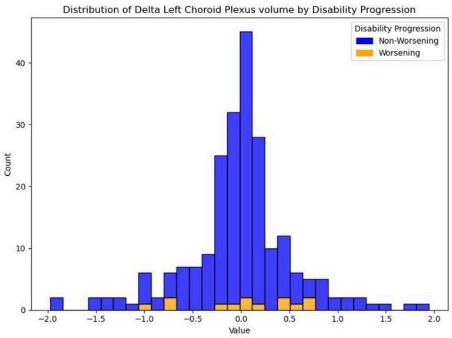
Distribution of Delta Left Choroid Plexus volume by Disability Progression
Disability Progression
Non-Worsening
Worsening
Count
Value

#### Appendix M: Left Lateral Ventricle Volume vs. Baseline Disability Score (EDSS_T0) by Disability Progression Status.

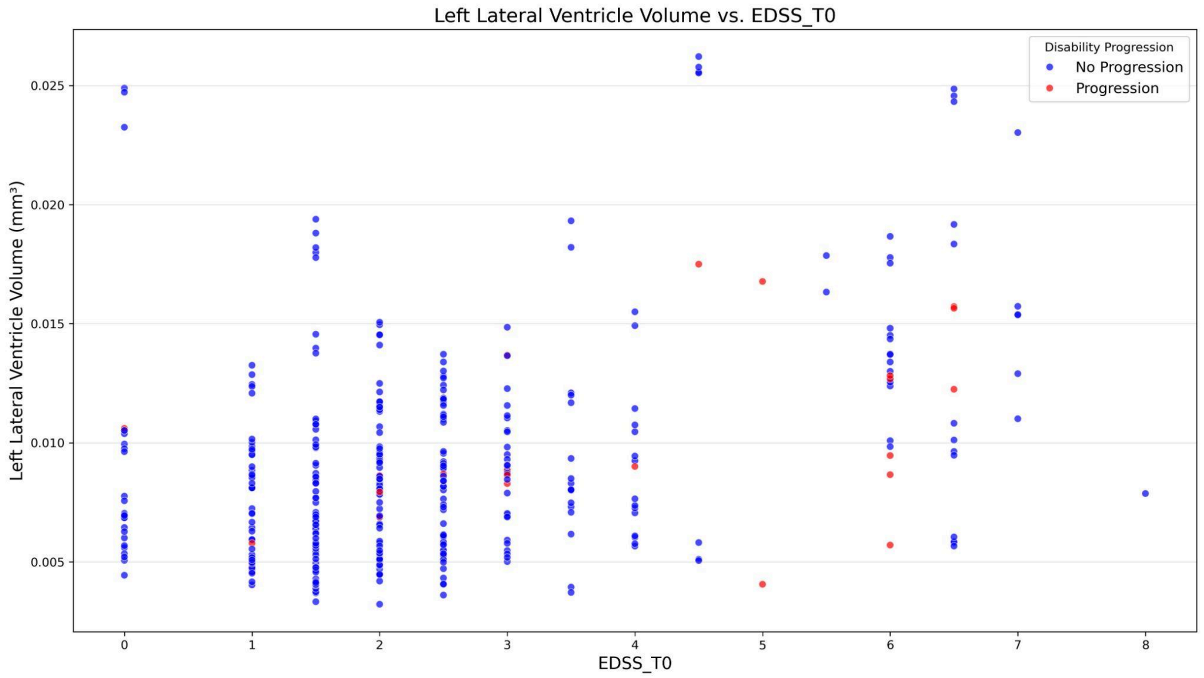

*Each point represents an individual case, colour-coded by progression status (blue for no progression, red for progression).*
